# Estimating Direct and Spillover Vaccine Effectiveness with Partial Interference under Test-Negative Design Sampling

**DOI:** 10.1101/2025.02.24.25322826

**Authors:** Cong Jiang, Fei Fang, Denis Talbot, Mireille E Schnitzer

## Abstract

The Test-Negative Design (TND), which involves recruiting care-seeking individuals who meet predefined clinical case criteria, offers valid statistical inference for Vaccine Effectiveness (VE) using data collected through passive surveillance, making it cost-efficient and timely. Infectious disease epidemiology often involves interference, where the treatment and/or outcome of one individual can affect the outcomes of others, rendering standard causal estimands ill-defined; ignoring such interference can bias VE evaluation and lead to ineffective vaccination policies. This article addresses the estimation of causal estimands for VE in the presence of partial interference using TND samples. Partial interference means that the vaccination of units within the same group/cluster may influence the outcomes of other members of the cluster. We define the population direct, spillover, total, and overall effects using the geometric risk ratio, which are identifiable under TND sampling. We investigate various stochastic policies for vaccine allocation in a counterfactual scenario, and identify policy-relevant VE causal estimands. We propose inverse-probability weighted (IPW) estimators for estimating the policy-relevant VE causal estimands with partial interference under the TND, and explore the statistical properties of these estimators.

## 1 Introduction

Vaccination campaigns targeting infectious diseases in populations have multifaceted effects, including boosting individual immunity and strengthening herd immunity Anderson and May (1985); Randolph and Barreiro (2020); Halloran and Hudgens (2018). Herd immunity, the indirect protection against infectious diseases, occurs when a large percentage of a population becomes immune through vaccination or prior infections.

Throughout the COVID-19 pandemic, the protective effects of mRNA vaccines have been evaluated using various methods, with many studies employing the test-negative design (TND) to assess real-world effectiveness (Dean et al., 2021; Schnitzer, 2022; Li et al., 2023). The TND is an observational study design that involves enrolling individuals exhibiting symptoms characteristic of the target infectious disease (such as COVID-19) who seek care for their symptoms and receive a test to determine the pathogen of interest (SARS-CoV-2, the virus that causes COVID-19) (Jackson and Nelson, 2013; Sullivan et al., 2014). The subsequent analysis compares the vaccination statuses between those who test positive (cases) and those who test negative (controls). An adjusted odds-ratio analysis produces an estimate of vaccine effectiveness (VE) for the prevention of outcomes of the target disease requiring care. Utilizing TND offers significant advantages in terms of efficiency and cost-effectiveness, while specifically helping to control for unobserved confounding differences in *healthcare-seeking behavior* between vaccinated and unvaccinated individuals Sullivan et al. (2014, 2016). However, the most causal interpretation of existing TND analytical strategies relies on the strong assumption that one person’s vaccination status does not impact another’s disease outcome, referred to as the “no interference” or “stable unit-treatment value (SUTVA)” assumption Cox (1958); Rubin (1980, 1986); Schnitzer et al. (2021). While this assumption may hold in certain contexts, it is not reasonable in many real-world applications, particularly in the context of vaccines and infectious diseases. For example, in a densely populated community, an individual’s vaccination status could potentially reduce the likelihood of transmission to others, thereby impacting their disease outcomes as well Ogburn and VanderWeele (2014). Due to this methodological limitation, it is necessary to consider the distinct effects of a vaccine on individual immunity versus its role in developing herd immunity.

Partial interference (Sobel, 2006) is the most methodologically studied type of interference, where study individuals are grouped into non-overlapping clusters (e.g., households, schools), with interference assumed to only occur between individuals within the same cluster. The methodological development of IPW estimators under partial interference for direct and spillover effects involves defining causal effect estimands (Hudgens and Halloran (2008)) and constructing corresponding estimators, with derivation of their asymptotic properties as the number of clusters grows (Tchetgen and VanderWeele, 2012; Perez-Heydrich et al., 2014; Liu et al., 2016; Barkley et al., 2020). Unlike traditional settings where the no interference assumption holds, partial interference necessitates knowledge of the entire probability distribution of treatment assignments within each cluster, known as the *cluster-level propensity score* (CPS), rather than just conditional expectations. Building on the foundational work of Tchetgen and VanderWeele (2012) and Liu et al. (2016), these IPW estimators allow unbiased estimation of average direct and spillover effects, accounting for intra-cluster interference. For inference, large sample (clusters) variance estimators from previous studies such as Perez-Heydrich et al. (2014) are utilized, and for scenarios with a small number of observed clusters, a bootstrap approach is recommended for more reliable inference Papadogeorgou et al. (2019). A notable application of the IPW method under partial interference is the evaluation of cholera vaccine effectiveness in a placebo-controlled, individually randomized trial in Matlab, Bangladesh Ali et al. (2005), which highlighted a significant spillover effect of cholera vaccination.

In observational settings, significant advancements have recently been made in causal estimation under the framework of partial interference. This includes developments such as parametric doubly robust estimators (Liu et al. (2019)), efficient semi-parametric estimation (Park and Kang (2022)), and efficient nonparametric sample-splitting estimators of stochastic policy effects (Lee et al. (2024)). The literature is particularly focused on *policy-relevant causal estimands*, which are defined under a counterfactual policy that modifies the distribution of treatment within the clusters Lee et al. (2024). These policies can be categorized into three types: (1) cluster-level proportions-based policies, including the Type-B policy (Tchetgen and VanderWeele (2012)) and generalized linear mixed model policies (GLMM, Papadogeorgou et al. (2019); Barkley et al. (2020)), (2) shifted propensity score policies, comprising the cluster incremental propensity score policy (CIPS, Kennedy (2019); Lee et al. (2024)) and cluster multiplicative shift policy (CMS, Wen et al. (2023); Lee et al. (2024)), and (3) the treated proportion bound (TPB) policy (Lee et al. (2024)). These hypothetical policies are more closely related to real-world interventions that modify recommendations and environmental factors (e.g., media, access to vaccination) rather than directly intervening on individual vaccination statuses. However, no work has yet proposed methodologies to evaluate direct, spillover, and overall effects of vaccination in the presence of partial interference under the TND, particularly in examining the different policies for these causal effects.

In this work, we focus on the estimation of policy-relevant causal estimands under partial interference from TND-sampled data, in the motivating context of mRNA vaccines to prevent COVID-19 outcomes. Existing partial interference methodological frameworks typically assume the random samples are taken from a super- or finite-population of clusters, with all members of the sampled clusters being subsequently observed. However, given the test-negative design sampling mechanism, we assume a super- or finite-population consisting of clusters of units, where only individuals within each cluster who meet the TND criteria are observed. To allow identifiability under the TND and unlike previous work, we propose defining the population direct effect (DE) and spillover effect (SE) using the geometric mean of risk ratios (RR). The geometric mean of RR not only appropriately reflects the multiplicative nature of risk ratios but also represents the “central tendency” on a logarithmic scale. This is consistent with the log-transformation approach, which stabilizes variance and normalizes the data distribution, thereby making statistical analysis more robust. Additionally, because of the form of the geometric mean of RR, corresponding DE and SE estimators can account for the varying sampling inclusion probabilities across different clusters, influenced by the unique characteristics of each cluster.

Our work considers VE estimation under the test-negative design (TND) in the presence of partial interference, where vaccination within a cluster may influence the outcomes of others. We define policy-relevant VE estimands using the geometric mean of risk ratios and propose inverse-probability weighted (IPW) estimators for their estimation. In Section 2, we introduce the TND sampling framework, define stochastic vaccine allocation policies, and establish the identifiability of direct and spillover effects under TND sampling. Section 3 presents our proposed IPW estimators, deriving their large-sample properties. In addition, building on M-estimation theory, we provide asymptotic results, demonstrating that our estimators are consistent and asymptotically normal under the correct specification of the cluster-level propensity score model. Lastly, Section 4 concludes the paper.

## 2 Methodology

### 2.1 Test-Negative Design Sampling and Notation

Suppose there are *m* observed clusters in the superpopulation according to the distribution *F*_0_. For each cluster *i* where *i* ∈ {1, …, *m*}, let *N*_*i*_ denote the total number of individuals in the underlying source population. In this paper, we consider the asymptotic region where the cluster size *N*_*i*_ = *O*(1) and the number of clusters *m* → ∞. We consider a test-negative design (TND) study that samples individuals hospitalized for COVID-19-like illness. For subject *j* = 1, …, *N*_*i*_ in cluster *i, V*_*i j*_ (categorical or binary) denotes COVID-19 vaccination status, *C*_*i j*_ represents measured confounders such as age, comorbidities, and employment sector, and *U*_*i j*_ represents unmeasured variables, which are assumed not to affect *V*_*i j*_. The infection statuses are represented by two indicator variables: 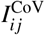, indicating SARS-CoV-2 infection, and 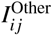, which equals 1 for other infections causing COVID-19-like symptoms. *W*_*i j*_ indicates COVID-19-like symptoms, occurring only if 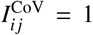 or 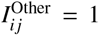, and *H*_*i j*_ indicates hospitalization for these symptoms. The TND samples from those with an infection 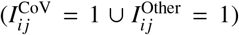, symptoms (*W*_*i j*_ = 1), and hospitalization (*H*_*i j*_ = 1). We use the indicator function 𝕀 to define 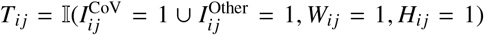, representing the inclusion criteria for the TND study. Further, by design, the set of infected and symptomatic individuals for whom *W*_*i j*_ = 1 and *H*_*i j*_ = 1 is the same as the set where *H*_*i j*_ = 1; therefore, eligibility for TND selection (*T*_*i j*_) is determined solely by hospitalization status (*H*_*i j*_), such that *T*_*i j*_ = *H*_*i j*_. Within each cluster, we observe censored samples with data 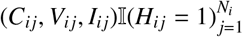 for *i* = 1, …, *m*. The combined outcome of interest is defined as 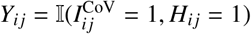, indicating a hospitalized (or medically-attended) symptomatic SARS-CoV-2 infection. This outcome involves three consecutive steps: infection with SARS-CoV-2, development of symptoms, and subsequent hospitalization due to these symptoms Schnitzer (2022). The event *Y*_*i j*_ = 1 identifies cases within the TND sample, while the condition {*Y*_*i j*_ = 0, *H*_*i j*_ = 1}, or equivalently 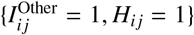}, corresponds to the controls in the sample.

### 2.2 Stochastic Policies

We define a *stochastic policy Q* ∼| **C**_*i*_, *N*_*i*_ as a probability distribution on 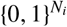 such that a cluster of size *N*_*i*_ with cluster-level covariate **C**_*i*_ receives vaccine ***v***_*i*_ with probability *Q* ***v***_*i*_ | **C**_*i*_, *N*_*i*_ in the conterfactual scenario. Previously proposed policies include the Type B policy (Tchetgen and VanderWeele (2012)), the generalized linear mixed-effects model (GLMM) shift policy (Papadogeorgou et al. (2019); Barkley et al. (2020)), cluster incremental propensity score policy (Kennedy (2019); Lee et al. (2024)), and cluster multiplicative shift policy (Wen et al. (2023); Lee et al. (2024)). First, the Type B policy (*Q*_B_) is defined as:

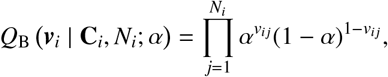

where α represents the independent and fixed probability of individuals receiving a vaccine, i.e., π_*i j*_ := P(*V*_*i j*_ = 1) = α for any individual *j* within cluster *i*. We note that under the Type-B policy, α denotes both the individual’s propensity and the cluster-average propensity to be vaccinated because the cluster-average propensity of treatment equals 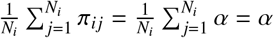.

In real-world settings, the relevance and practicality of the Type B policy are limited; therefore, Papadogeorgou et al. (2019) and Barkley et al. (2020) suggested alternatives wherein vaccination probabilities may vary among individuals. The GLMM shift policy of Papadogeorgou et al. (2019) allowed for the individual’s propensity to depend on covariates, while ensuring that the cluster-average propensity of treatment remains at a certain hypothetical value (say β). Specifically, for an individual *j* within cluster *i* and with covariate vector *C*_*i j*_, the probability of receiving the vaccine can be represented as 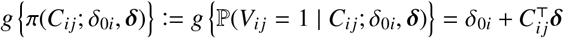 using the appropriate link function *g*, for some fixed, pre-specified value of *δ*, and for δ_0*i*_ satisfying 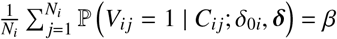. Then, in the counterfactual world, under the assumption that the variables 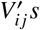 are conditionally independent given **C**_*i*_, and *V*_*i j*_ is conditionally independent of **X**_*i*,− *j*_ given *C*_*i j*_, the corresponding GLMM policy (denoted as *Q*_GLMM_) is defined as

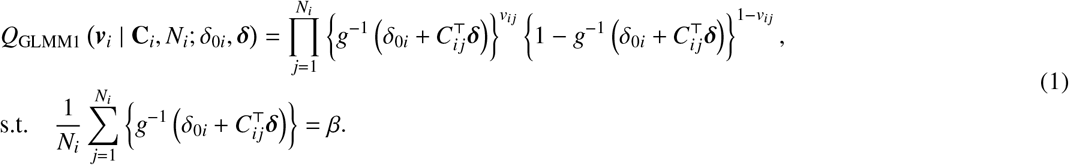

Note that, for each cluster, the cluster-average propensity of treatment is set at a certain hypothetical value β. Alternatively, one may consider including cluster-specific random effects in the policy formulation. Incorporating a cluster-specific random effect *u*_*i*_ assumed to follow a distribution, e.g., a Gaussian distribution function Φ with mean zero and variance σ^2^, the probability of an individual *j* within a cluster *i* receiving the vaccine is 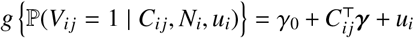 with a constant *γ*_0_; then, the second type GLMM policy can be defined as

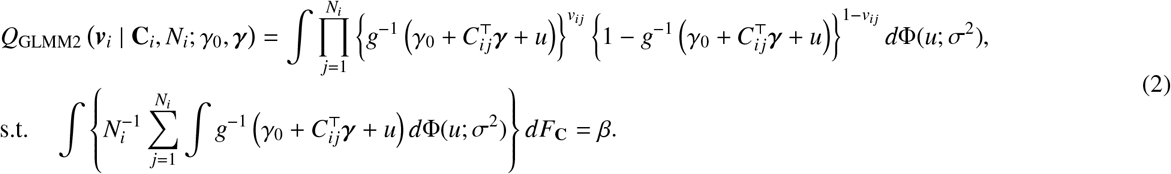

Under a certain assumption discussed in Barkley et al. (2020) Section 4.2, i.e., the conditional odds ratio of treatment remains identical for any two individuals within a cluster, regardless of whether they are in the factual or counterfactual scenario, the parameters *γ* and those in the distribution Φ can be identifiable from the observed treatment GLMM model. Unlike the previously defined *Q*_GLMM1_ in Eq. (1), where *V*_*i j*_s are conditionally independent, the *Q*_GLMM2_ in Eq. (2) accounts for correlation among individuals within the same cluster because of the cluster-specific random effects, with the degree of correlation depending on the variance of Φ. Moreover, different from *Q*_GLMM1_, which sets the cluster-average propensity of treatment to β, *Q*_GLMM2_ allows for greater flexibility by ensuring that the marginal probability of treatment selection equals β, while the average individual-level propensity scores within any given cluster may deviate from β.

Building upon Kennedy (2019)’s incremental propensity score interventions for the context of clustered interference, Lee et al. (2024) introduced the Cluster Incremental Propensity Score (CIPS) policy. This policy shifts the observed propensity score distribution by a user-defined function of **C**_*i*_ and *N*_*i*_, denoted as *γ*(**C**_*i*_, *N*_*i*_). Specifically, the counterfactual treatment odds are *γ*(**C**_*i*_, *N*_*i*_) times the observed odds. Formally, for the propensity score (π_*i j*_) of individual *j* in cluster *i*, defined as π_*i j*_ = P(*V*_*i j*_ = 1 | **C**_*i*_, *N*_*i*_), the shifted propensity score 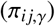 is given by 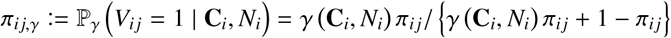, ensuring that the odds ratio 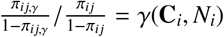.

The CIPS policy can then be expressed as

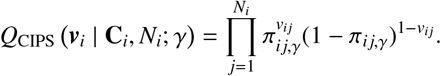

Notably, the CIPS policy preserves the within-cluster ranking of unit vaccination probabilities without relying on parametric models. Varying the *γ* function allows for the generation of different impacts of the policy on the propensity scores. For instance, setting *γ*(**C**_*i*_, *N*_*i*_) to a constant value of 2 results in a scenario where we investigate the relative risk of COVID-19 under the policy that doubles the current odds of vaccination.

A recently defined policy known as the cluster multiplicative shift (CMS) policy, which extends the multiplicative shift policy of Wen et al. (2023) to the partial interference setting, was investigated in Lee et al. (2024). CMS selectively increases the vaccination propensity for individuals with specific covariate values or patterns, such as those indicating individuals at high risk for adverse outcomes. We can use this kind of policy to estimate effects on mean outcomes under counterfactual scenarios where a larger proportion of high-risk individuals receive vaccination. For instance, a CMS policy can be defined based on a shifted propensity score, determined by a user-specified factor λ, and some single-dimensional binary covariate 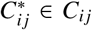, such that 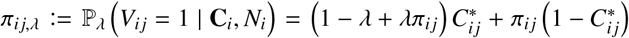, representing the shifted probability of treatment for individual *j* in cluster *i*. Note that the propensity score distribution is adjusted solely for individuals with 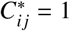, thereby preserving the conditional independence of treatment selection within clusters. The CMS policy can then be expressed as 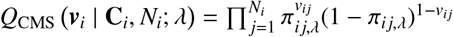.

### 2.3 Policy-relevant Causal Estimands and Identifiability

Under partial interference, for *m* clusters, each containing *N*_*i*_ individuals, where *i* ∈ {1, …, *m*}, we first define 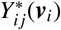 as the potential outcome for individual *j* in cluster *i* if the vaccination status of cluster *i* is *N*_*i*_-vector ***v***_*i*_. Then, we define the *individual average potential outcome* for individual *j* in cluster *i* under policy *Q* as:

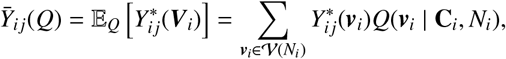

where the sum is taken over all possible configurations of cluster-level vaccine vectors ***v***_*i*_ under the policy distribution *Q*. This expression represents the expected outcome of individuals in the counterfactual scenario where the vaccine is allocated based on policy *Q*, averaging over all possible vaccination configurations within the cluster given the policy *Q*. Next, we define the *cluster average marginal potential outcome* for cluster *i* under policy *Q* by

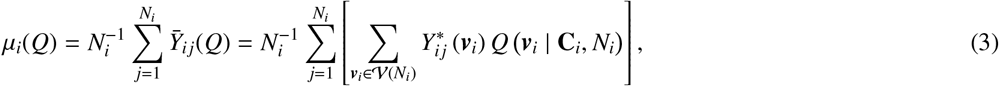

which takes the average over all the units in a cluster, considered as a finite population of a certain size (e.g., *N*_*i*_ for cluster *i*). Similarly, we define the *cluster average potential outcome* for cluster *i* under policy *Q* when a given individual *j* receives vaccine *v* by

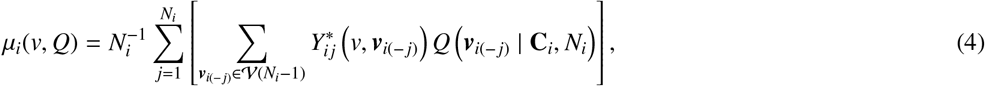

which represents the cluster *i*’s averaged outcome in the counterfactual world where the vaccine is assigned according to policy *Q*, but the vaccine of individual *j* is independently set to *v*.

To represent causal effects, we define the cluster-level policy-relevant causal estimand by contrasting µ_*i*_(*v, Q*) and µ_*i*_(*Q*) across different values of *v* and policies *Q* (Tchetgen and VanderWeele, 2012). *Varying either the value of v* or the policy *Q* while holding the other fixed, we define the *population direct effect* and *population spillover effect* based on the definition of the geometric risk ratio as follows.

#### Definition 1

*(Population direct and spillover effects of the geometric risk ratio). Assuming that clusters are observed from a super-population according to a distribution F*_0_, *the geometric risk ratio describes the geometric mean of the risk ratios:*

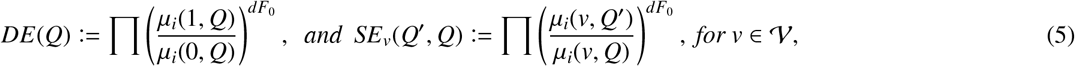

*where Π denotes the product operator if* **C**_*i*_ *is discrete, or the product integral if* **C**_*i*_ *is continuous*.

The above-defined population direct and spillover effects of the geometric risk ratio can also be expressed as follows:

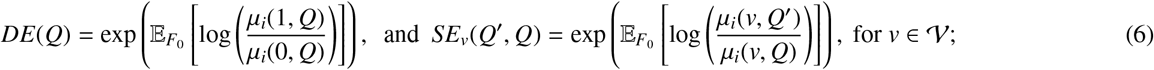

since, for DE, for example, we have

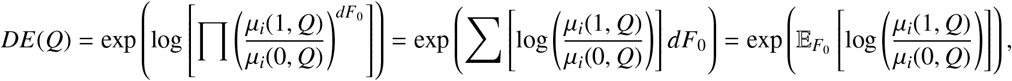

Then, the *VE direct effect* and *VE spillover effect* can be written as 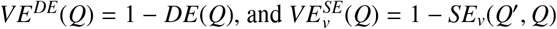, respectively. Following Definition 1, the proposed framework introduces a novel approach to quantifying the population direct effect and spillover effect using the geometric mean of risk ratios. This approach is particularly advantageous within the context of vaccine effectiveness, as it aligns well with the inherent multiplicative nature of risk ratios and accurately reflects the “central tendency” on a logarithmic scale. The geometric mean serves not only to stabilize variance and normalize the data distribution, enhancing the robustness of our statistical analysis, but also to address the challenges linked to varying sampling inclusion probabilities across different clusters under TND. By leveraging the risk ratio at the cluster level, this method effectively cancels out common but unknown or unquantifiable terms in the estimation process, such as those inherent in 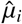. The subsequent section 3.2.1 will delve deeper into the mechanics and rationale behind TND sampling, offering comprehensive insights into how the geometric mean framework contributes to more accurate and interpretable measures of the direct and spillover effects within diverse cluster settings.

Comparing to the previous work defined, e.g., the direct effect, using a ratio scale as the form of

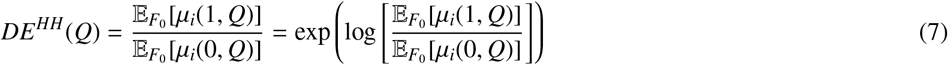

where the superscript *HH* in *DE*^*HH*^ represents the notions of direct, indirect (spillover), total, and overall causal effects as defined by Hudgens and Halloran (2008). The difference between our estimand and their estimand can be characterized as follows.

#### Lemma 1

**(Difference between** *DE*(*Q*) **and** *DE*^*HH*^(*Q*)) *Let DE*(*Q*) *and DE*^*HH*^(*Q*) *be defined as in equations* (6) *and* (7), *respectively. Then*

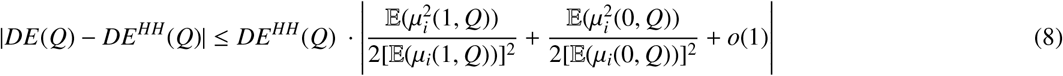

Similar bounds apply to *S E*^*HH*^(*Q, Q*^′^), *TE*^*HH*^(*Q*), and *OE*^*HH*^(*Q*) by replacing µ_*i*_(·)) with the corresponding quantities in each estimand. Lemma 1 shows that the difference between *DE*(*Q*) and *DE*^*HH*^(*Q*) is of order one and should therefore be expected. Moreover, if both *DE*(*Q*) and the terms on the right-hand side of the inequality can be consistently estimated, then this result provides a basis for constructing upper and lower bounds for *DE*^*HH*^(*Q*) using an estimator of *DE*(*Q*).

Because log(*x*) is a strictly convex function of *x*, we have, using Jensen’s inequality,

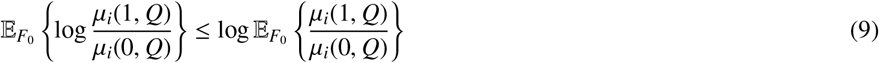

and the proposed *DE*(*Q*) ≤ *DE*^*HH*^(*Q*).

Based on the above definitions of direct and spillover effects, we can further define the *population total effect* and *population overall effect* using the geometric risk ratio as follows:

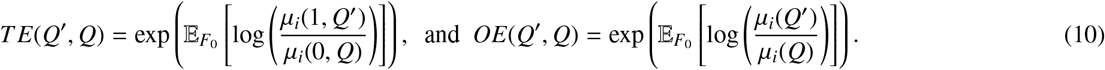

do we need this paragraph? It follows immediately from their definitions that the total effect can be decomposed as the product of direct and indirect causal effects, such that *TE*(*Q*^′^, *Q*) = *DE*(*Q*^′^)*SE*_0_(*Q*^′^, *Q*). The VE total effect *VE*^*T E*^(*Q*^′^, *Q*) = 1 − *TE*(*Q*^′^, *Q*) measures the reduction in the risk of infection for a vaccinated individual in a cluster assigned treatment based on one policy *Q*^′^ compared to a counterfactual scenario where the unvaccinated individual is in the cluster from the same study population assigned treatment based on another policy *Q*. The VE overall effect *VE*^*OE*^(*Q*^′^, *Q*) = 1 − *OE*(*Q*^′^, *Q*) measures the reduction in the risk of infection for an individual, whether vaccinated or unvaccinated, in a cluster with treatment assignment based on one policy *Q*^′^ compared to what would have happened to an individual in the cluster from the same study population assigned treatment based on another policy *Q*.

The standard identification of direct, spillover, total and overall effects requires three assumptions: (1) *Consistency:*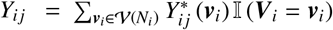; (2) *Conditional Exchangeability:* For 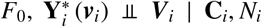 for all ***v***_*i*_ ∈ 𝒱 (*N*_*i*_) ; (3) *Positivity:* for some ϵ ∈ (0, 1), ϵ < ℙ ***V***_*i*_ = ***v***_*i*_ | **C**_*i*_, *N*_*i*_ < 1 − ϵ for all ***v***_*i*_ ∈ 𝒱 (*N*_*i*_), and applying iterated expectations, the causal estimands defined in Equaitons 4 and 3 can be identified as

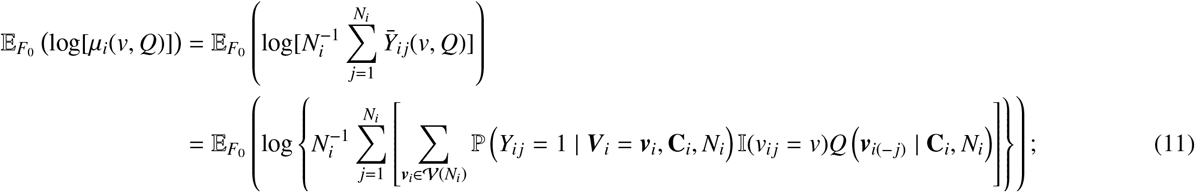

And

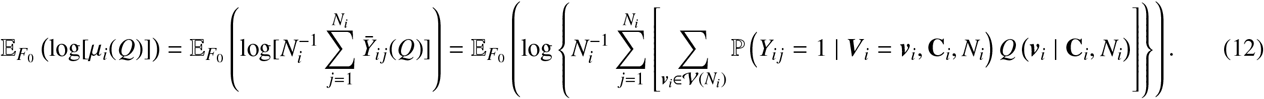

See detailed proof in Appendix A. Building on the above two identified policy-relevant causal estimands, the defined population direct, spillover, total, and overall effects of the geometric risk ratio can be correspondingly identified; for example, for the population direct effects, we have:

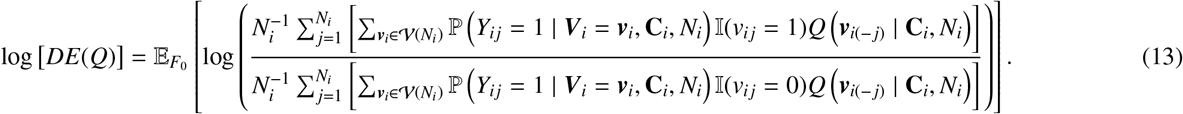

## 3 Estimation of Policy-relevant Estimands

### 3.1 Estimation under simple random sampling (SRS)

For each cluster, consider observed data under simple random sampling (SRS) from a finite population (with size *N*_*i*_) of cluster *i* denoted by *Y*_*ik*_, *C*_*ik*_, *V*_*ik*_, *k* ∈ 𝒮_*i*_, *i* = 1, 2, …, *M*. For example, the cluster sample mean of the outcome of cluster *i* is written 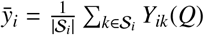, where |𝒮_*i*_| is the cluster sample size of the simple random sample 𝒮_*i*_.

In causal inference, the propensity score (PS) (Rosenbaum and Rubin, 1983) is used to balance the distributions of confounders between the treatment and the control group. In the presence of partial interference, the cluster-level PS, *τ* (***v***_*i*_ | **C**_*i*_, *N*_*i*_) = ℙ ***V***_*i*_ = ***v***_*i*_ | **C**_*i*_, *N*_*i*_, is the joint probability of the treatment status of all individuals within the cluster given all individuals’ covariates (Zhang et al. (2023); Kang et al. (2023)).

Under SRS within each cluster, for *v* ∈ 𝒱, inverse probability weighting (IPW) estimators of µ_*i*_(*v, Q*) and µ_*i*_(*Q*) are

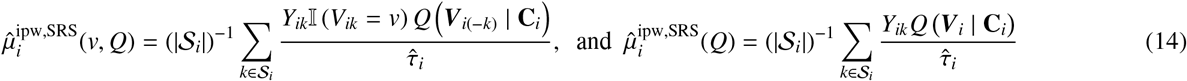

where, with parameters *α* and 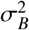, the cluster-level PS, specifically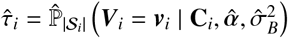, is modelled as follows. Building upon the previous work of Tchetgen and VanderWeele (2012), Perez-Heydrich et al. (2014), Liu et al. (2016), Barkley et al. (2020), and Kang et al. (2023), we employ a mixed-effects model for the treatment to model the cluster-level PS, with the link function *g* (e.g., the logit link):

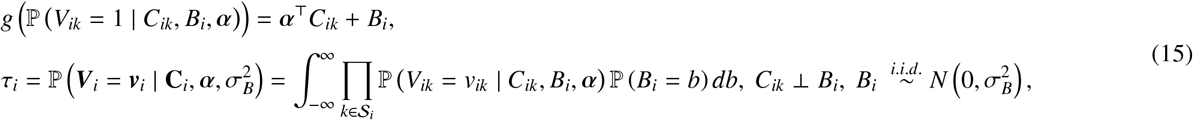

where the Normal random effect *B*_*i*_ with mean 0 and variance 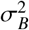 accounts for the correlation in treatment selection among individuals within the same cluster. Then, the model parameters for the cluster-level PS, which includes both *α* and 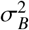, are estimated by 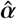 and 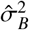 using maximum likelihood estimation or a restricted maximum likelihood estimation (Bates et al., 2015).

To demonstrate the unbiasedness of the estimators for the cluster average marginal potential outcome for *v* ∈ 𝒱 and the cluster average potential outcome, we consider the randomness arising from the simple random sampling and, conditional on the sampling, the randomness from the treatment assignments. Specifically, we have:

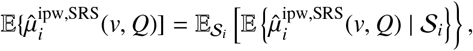

where the outer expectation 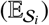 is with respect to the probability sampling design specified by the probability measure 𝒫 over the set of all possible candidate samples. For simple random sampling without replacement of cluster *i*, 𝒫(𝒮_*i*_) is given by 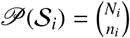 if |𝒮_*i*_| = *n*_*i*_; otherwise, 𝒫(𝒮_*i*_) = 0, where, again, *N*_*i*_ is the total number of individuals in the underlying source population for cluster *i*, and *n*_*i*_ is the SRS sample size for cluster *i*.

For the inner expectation, following the theories outlined in existing IPW estimators for partial interference by Tchetgen and VanderWeele (2012), Perez-Heydrich et al. (2014), or Papadogeorgou et al. (2019), we can demonstrate that the estimators in Equation (14) are unbiased for the cluster average potential outcome if the cluster-level propensity score model (15) is correctly specified. Consequently, the corresponding population total effect estimator (as well as those of the other defined effects), which relies on Equation (14), is consistent and asymptotically normal, and the sandwich-type variance estimators are similar to those in Perez-Heydrich et al. (2014). The detailed results under SRS are given in Appendix D for our proposed population direct and spillover effects of the geometric risk ratio. Besides the above results regarding the SRS of each cluster in the partial interference analysis, the goal of this subsection is to introduce sampling randomness into our analysis, which leads to our focus on TND sampling within clusters.

### 3.2 Estimation under the test-negative design sampling

#### 3.2.1 TND sampling mechanism in non-probability sampling and assumptions

The TND is a type of non-probability sampling. Unlike probability sampling, where each element of the population has a known chance of being selected, non-probability sampling includes methods where some population elements have no chance of selection or their selection probability is not known in advance (Wu and Thompson (2020)). Thus, one typically selects and fits a model for the unknown selection mechanism, where the outcome is the inclusion indicator variable based on the unit’s characteristics. However, in TND sampling, the selection probability cannot be estimated because we only have information on the selected individuals and not on the source population, which is nevertheless our target of inference. Fortunately, the risk ratio form of the target estimand helps us circumvent the issue of the inestimable selection probability (Schnitzer et al., 2021).

Consider the inclusion criterion for the test-negative design. Let the TND-sampled data for cluster *i* be denoted as *k* ∈ 𝒮_TND,*i*_, where *i* = 1, 2, …, *M*. Define the TND sample indicator variable *H*_*i j*_ as 𝕀 (*j* ∈ 𝒮_TND,*i*_) for *j* = 1, 2, …, *N*_*i*_, with the indicator function 𝕀. This is a Bernoulli random variable under the TND sampling design and is defined for every *j* in the cluster *i*, with probability 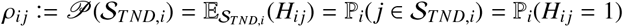, where 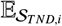 is the expectation with respect to the TND sampling design over the set of all possible candidate TND samples. Then, building on the Horvitz-Thompson principle, for *j* = 1, 2, …, *N*_*i*_, and any fixed function *f* we have the TND sampling unbiasedness equality, which we refer to as the *Standard TND Sampling Debiased Principle*, as follows:

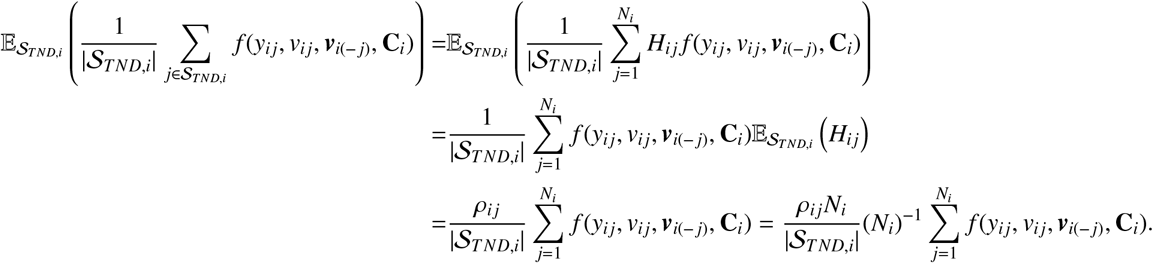

It is important to note that the factor term in the TND Sampling Debiased Principle, i.e., *ρ*_*i j*_*N*_*i*_ |𝒮_*T ND,i*_|, includes elements *ρ*_*i j*_ and *N*_*i*_, which is unknown and cannot be estimated using TND samples. In addition, in the presence of interference, dependence between individuals’ *H* is another layer of the complicity of the problem, due the the casual path such that 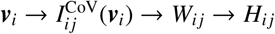.

However, in the presence of partial interference, the standard identifiability condition based on the inverse of individual-level selection probabilities (e.g., 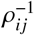) in test-negative design (TND) studies is not necessary. Instead, it becomes essential to adopt a more appropriate quantity that captures the bias induced by the TND selection mechanism under interference. To address this, we introduce the *joint TND inclusion probability* for each cluster, defined as, with 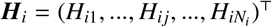 the TND inclusion indicator vector,

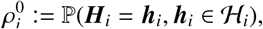

where ℋ_*i*_ denotes the set of valid inclusion patterns for the TND sample in cluster *i*, i.e.,

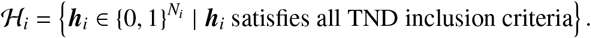

Unlike the marginal inclusion probability *q*_0_ = ℙ (*H* = 1) assumed in non-interference settings, 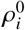 captures within-cluster dependencies, including potential correlations between individual outcomes and their peers’ selection statuses. This refinement allows for a more accurate representation of the distribution shift between the observed TND sample and the target population, supporting valid inference under partial interference. Our target estimand is expressed on the risk ratio scale, which has been advocated in recent work (e.g., Schnitzer (2022), Jiang et al. (2023)) as a robust choice under such settings.

Moreover, our newly defined population-level direct and spillover effects based on the geometric risk ratio, which aggregate cluster-specific risk ratios, allow for the cancellation of inestimable terms. In particular, the joint TND selection probabilities 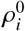 may vary across clusters due to differences in realized values of ***h***_*i*_, resulting in heterogeneity in 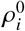. As a consequence, the form of *DE*^*HH*^(*Q*) is only *partially identifiable*, since these heterogeneous selection probabilities cannot be canceled out across clusters. In contrast, the geometric formulation defined in Definition 1 enables cancellation of the selection probabilities within each cluster, rendering the estimands fully identifiable. See Lemma 5 for details.

Finally, we present three assumptions under TND sampling to ensure valid statistical estimation. For the cluster *i*, with ***H***_*i*_ the vector of TND selection indicators for individuals within cluster *i* and ℋ_*i*_ denotes the set of inclusion patterns that satisfy all TND criteria, we assume (A1) ℙ_*i*_(**Y**_*i*_ = **y**_*i*_ | ***V***_*i*_, **C**_*i*_, ***H***_*i*_ = ***h***_*i*_, ***h***_*i*_ ∈ ℋ_*i*_) = 0 for **y**_*i*_ ∈ {**0, 1**}; (A2) 0 < ℙ_*i*_ ***V***_*i*_ = ***v***_*i*_ | **C**_*i*_, ***H***_*i*_ = ***h***_*i*_, ***h***_*i*_ ∈ ℋ_*i*_ < 1; (A3) 0 < ℙ_*i*_(**C**_*i*_ = **c**_*i*_ | ***H***_*i*_ = ***h***_*i*_, ***h***_*i*_ ∈ ℋ_*i*_) < 1; (A4) cluster-level control exchangeability assumption: in each cluster, being hospitalized for symptoms of *another infection* is independent of vaccination conditional on covariates, that is {**Y**_*i*_ = **0, *H***_*i*_ = ***h***_*i*_, ***h***_*i*_ ∈ ℋ_*i*_} ⫫ ***V***_*i*_ | **C**_*i*_.

(A1) - (A3) are essentially positivity-type cluster-level assumptions for the TND samples in each cluster. Specifically, (A1) requires that each cluster contains both cases and controls. (A2) requires positivity for the treatment assignment and (A3) pertains positivity for the cluster-level covariates. Lastly, (A4) is the cluster-level control exchangeability assumption that was discussed in Jiang et al. (2023), which is sufficient but not necessary for the typical TND assumption that the vaccine does not have an impact on non-target infections (Feng et al., 2017).

#### 3.2.2 Proposed estimation under the TND sampling

Under cluster-level TND sampling, with the sample data for cluster *i* and *k* ∈ 𝒮_*T ND,i*_ for *i* = 1, 2, …, *m*, we have the following inverse probability weighting estimators:

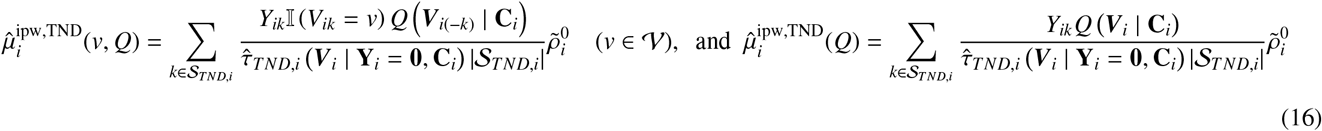

where |𝒮_*T ND,i*_| is the cluster sample size of the TND sample 𝒮_*T ND,i*_, and 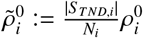 where 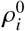 is the joint TND inclusion probability of cluster *i*, and

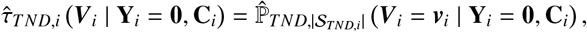

which is the cluster-level propensity score, estimated from control data in TND samples (i.e., {**Y**_*i*_ = **0, *T***_*i*_ = **1**}). Further, similar to the estimation of the cluster propensity score under the SRS sampling in Equation (15), we have model that

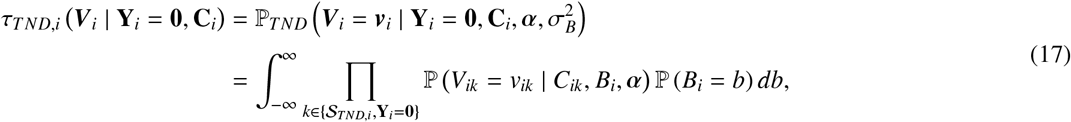

Where 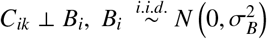, and 𝕡 (*V*_*ik*_ = 1 | *C*_*ik*_, *B*_*i*_, *α*) = *g*^−1^ (*α*^⊤^*C*_*ik*_ + *B*_*i*_). To estimate *α* and 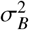, we maximize the log-likelihood for the mixed effects model

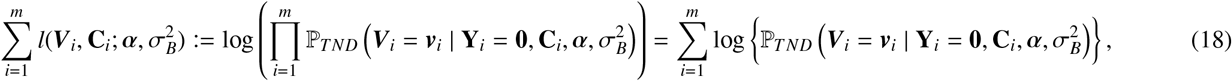

with respect to *α* and 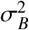, yielding 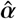 and 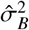.

##### Lemma 2

*(Identifiability for cluster-level PS) Under cluster-level control exchangeability assumption, i*.*e*., {**Y**_*i*_ = **0, *H***_*i*_ = ***h***_*i*_, ***h***_*i*_ ∈ ℋ_*i*_} ⫫ ***V***_*i*_ | **C**_*i*_, *then the cluster-level propensity score τ*_*i*_ = ℙ ***V***_*i*_ = ***v***_*i*_ | **C**_*i*_ *can be identified as τ*_*T ND,i*_ ***V***_*i*_ | **Y**_*i*_ = **0, C**_*i*_ = ℙ_*T ND*_ ***V***_*i*_ = ***v***_*i*_ | **Y**_*i*_ = **0, C**_*i*_.

##### Proof

See Appendix B, Section 1.

##### Proposition 1

*(Consistent estimation for cluster-level PS) Assume that the cluster-level propensity model* 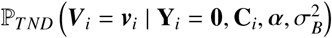 *is correctly specified such as in Equation (17)*. *If the parameters* 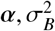 are estimated by the MLE estimators for the mixed effects model in Equation (18), then, under cluster-level control exchangeability assumption, the cluster-level PS *τ*_*i*_ = ℙ (*V*_*i j*_ = *v*, ***V***_*i*(− *j*)_ = ***v***_*i*(− *j*)_ | **C**_*i*_) *is consistently estimated by*

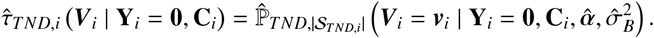

*In addition*, 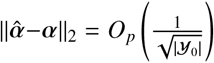, *where* |𝒴_0_| *is the total number of TND observed controls among all m clusters, and* 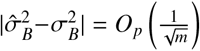. *Jiang (2007)*

Note that, unlike previous cluster-level propensity score (PS) estimators for inverse probability weighting (IPW) under partial inter-ference—such as those proposed in Tchetgen Tchetgen et al. (2009); Liu et al. (2016); Papadogeorgou et al. (2019)—our consistency result for the cluster-level PS estimator holds under both repeated test-negative design sampling within a cluster and repeated treatment assignments. In contrast, the consistency of earlier estimators applies only to repeated treatment assignments, assuming fixed cluster sizes.

##### Corollary 1

(Consistent estimation for *τ*_*T ND,i*_*) Let* 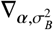 *denote the gradient of τ*_*T ND,i*_(**V**_*i*_ | **Y**_*i*_ = **0, C**_*i*_) *with respect to the p* + 1 *parameter vector* 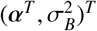. *Suppose there exists a constant C*_0_ > 0 *such that* 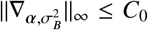. *Then, under the convergence rates for* 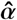 *and* 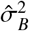 *established in Proposition 1, it holds with probability* 1 − *o*_*p*_(1) *that*

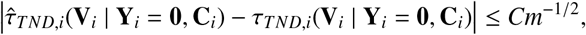

*for some constant C* > 0 *and i* ∈ [*m*].

**Proof 1** *From the formula in equation* (17) *and the differentiability of the inverse logistic function, we apply first-order Taylor expansion to obtain*

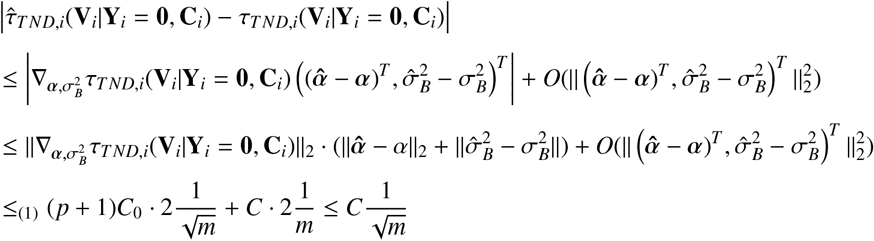

*where (1) follows from the Cauchy–Schwarz inequality, along with the boundedness of the gradient and the convergence rates established in Proposition 1*.

##### Theorem 1

*(Unbiasedness of IPW Estimators Based on TND Sampling) Let* 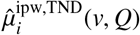 *and* 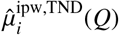 be the estimators defined in Equations (16) based on TND samples and if the cluster-level propensity score *τ*_*i*_ = P (***V***_*i*_ = ***v***_*i*_ | **C**_*i*_)*is known, then* 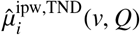 *and* 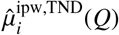 *are unbiased estimators of* µ_*i*_(*v, Q*) *and* µ_*i*_(*Q*), *respectively, as defined in Equations (11)*.

**Proof:** See Appendix B.

In addition, replacing |𝒮_*T ND i*_| with its consistent estimator 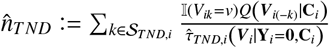 yields a Hájek-type estimator Hájek (1971); Liu et al. (2016). It is well established that the Hájek-type inverse probability weighting (IPW) estimator exhibits superior efficiency relative to the traditional IPW estimator in finite samples Särndal et al. (2003). Details regarding the construction of the Hájek-type IPW estimator are provided in the Appendix C.

Regarding the target direct and spillover effects in Equations (6), we propose the following estimators:

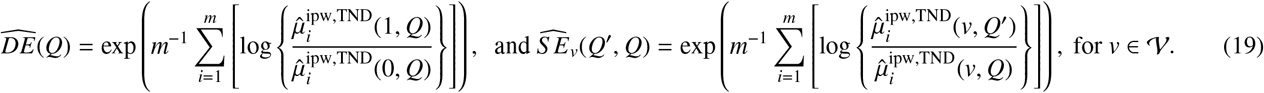

It is important to note that 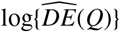 is a solution for log{*DE*(*Q*)} to the estimating equation

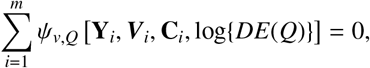

where

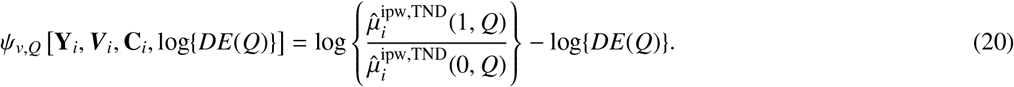

Further, for the score functions system of cluster-level propensity score Equation (18), we have the estimating equation that 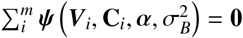, where

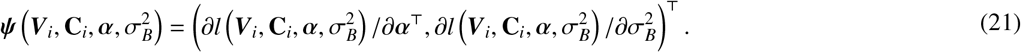

Building on these estimating equations, the following theorem addresses the asymptotic normality of 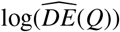:

##### Theorem 2

*(Asymptotic Normality of the Proposed IPW-type Estimators Based on TND Sampling) Assume that the parametric propensity score model is correctly specified and that the regularity assumptions in the Appendix hold. Then, the following results hold:*

1. 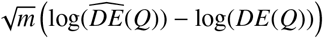 *converges in distribution to N*(0, Σ_*log*(*DE*)_), *as m* → ∞, *where* Σ_*log*(*DE*)_ *is expressed as*

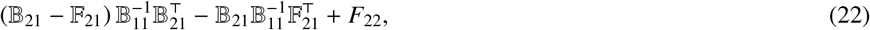

*where*

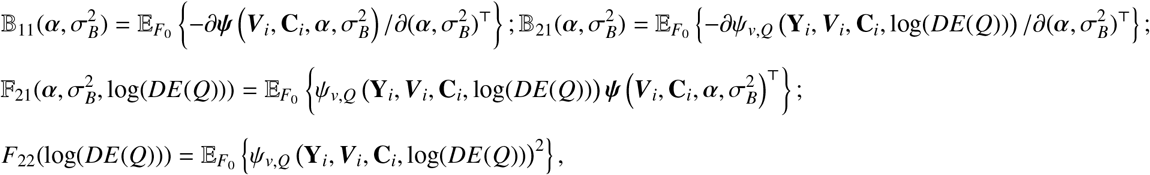

*regarding estimation equations in Equations 20 and 21*.

##### Proof

See Appendix D. Similar properties related to spillover, total, and overall effects are also elaborated in the same appendix.

In addition, by applying the Delta Method with the exponential function such that exp[log(*DE*(*Q*))] = *DE*(*Q*), we have that 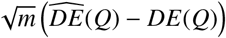 converges in distribution to *N*(0, Σ_*DE*_), as *m* → ∞, where Σ_*DE*_ = (*DE*(*Q*))^2^Σ_*log*(*DE*)_. The regularity conditions and detailed steps are verified in Appendix D, ensuring the application of the central limit theorem to our estimators. The asymptotic variances Σ_0_ and Σ_*T ND*_ are derived based on the specifics of the IPW-type estimators and the TND sampling framework.

##### Algorithm 1

Partial Interference Test-Negative Design (PI-TND) direct and indirect VE IPW Algorithm

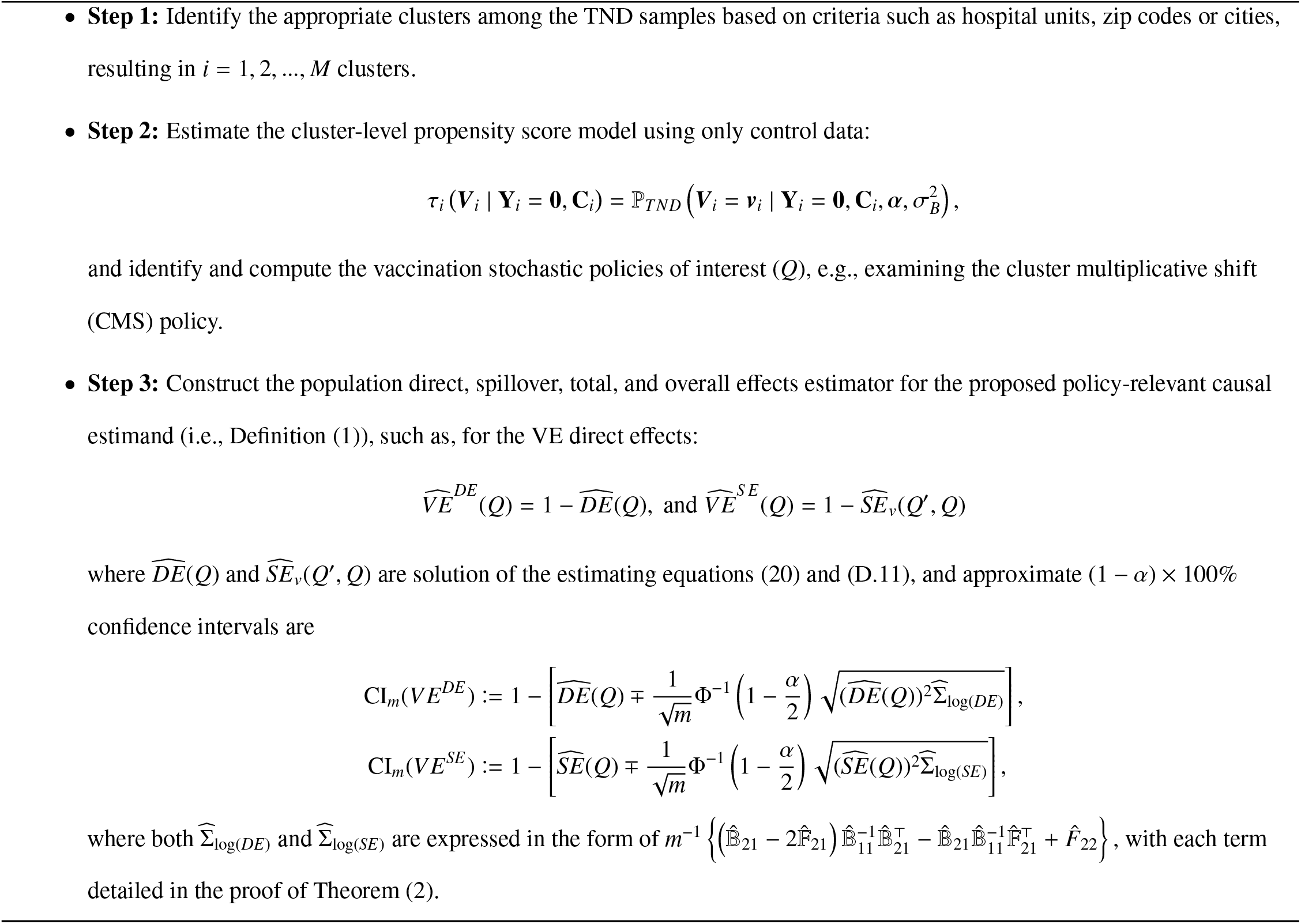

Thus, building on the above results and using cluster-level TND samples, our defined population direct, spillover, total, and overall effects, as well as the corresponding VE metrics, can be consistently estimated. We acknowledge that not only the biased term (the joint TND inclusion probability) but also the estimator 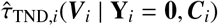 cancels out across clusters in the ratio 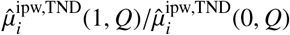. Nevertheless, its estimation remains essential, as it plays a critical role in assessing the overlap between observed and hypothetical treatment assignments, which in turn informs the scale of variance for the estimators (e.g., Σ_*log*(*DE*)_ in Equation (22)).

##### Lemma 3

*Suppose there exist constants* Γ_1_, Γ_2_ > 0 *such that* 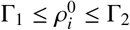 *for all i* ∈ [*m*]. *Then, with probability* 1 − *o*_*p*_(1),

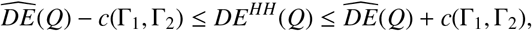

*Where*

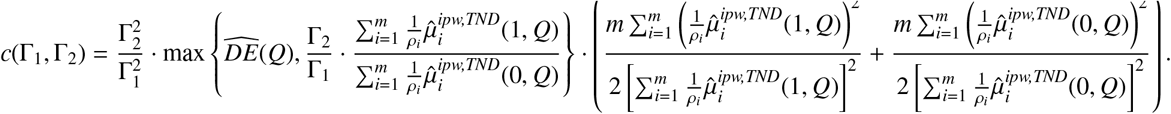

Note that the quantity 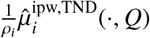 is fully identifiable from the TND sample. While the realized values of *H*_*i*_ may vary substantially across clusters, the identical distribution across clusters ensures that the lower and upper bounds Γ_1_ and Γ_2_ for 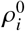 are the same across *i* ∈ [*m*].

Lemma 4 demonstrates that if prior knowledge about Γ_1_ and Γ_2_ is available, one can obtain an informative range for the magnitude of *DE*^*HH*^(*Q*). In practice, by considering multiple plausible values for (Γ_1_, Γ_2_), investigators can assess the sensitivity of *DE*^*HH*^(*Q*) to variation in these bounds. It is important to emphasize that the purpose of Lemma 4 is to elucidate the relationship between our proposed estimand and the corresponding estimand in the existing literature, rather than to facilitate inference for the estimator of *DE*^*HH*^(*Q*). Lemma 4 can be extended analogously to the spillover, total, and overall effects—namely *S E*^*HH*^(*Q, Q*^′^), *TE*^*HH*^(*Q*^′^, *Q*), and *OE*^*HH*^(*Q*^′^, *Q*)—by replacing 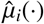 with the corresponding terms from the respective estimators of *S E*(*Q, Q*^′^), *TE*(*Q*^′^, *Q*), and *OE*(*Q*^′^, *Q*) respectively.

##### Lemma 4

**(Difference between** *DE*(*Q*) **and** *DE*^*HH*^(*Q*)) *Assume there exist constants* Γ_1_, Γ_2_ > 0 *such that* Γ_1_ ≤ *ρ*_*i*_ ≤ Γ_2_ *for all i* ∈ [*m*]. *Then, there exists a constant C* > 0 *such that, with probability* 1 − *o*_*p*_(1),

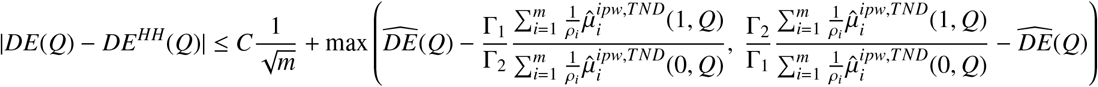

The bound on the right-hand side is estimable and facilitates sensitivity analysis through varying the constants (Γ_1_, Γ_2_). Notably, the estimands *DE*(*Q*) and *DE*^*HH*^(*Q*) do not preserve order under policy shifts in general. That is, for two policies *Q* and *Q*^′^, it is generally not the case that *DE*(*Q*) > *DE*(*Q*^′^) implies *DE*^*HH*^(*Q*) > *DE*^*HH*^(*Q*^′^).

**Proof 2** *The difference between DE*(*Q*) *and DE*^*HH*^(*Q*) *can be decomposed as*

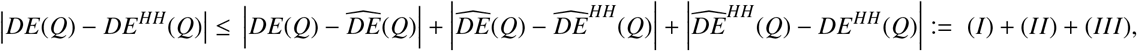

*where* 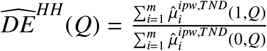 *is a partially identifiable plug-in estimator that depends on the unknown parameters *ρ*_*i*_. For term (I), applying the delta method to* log *DE*(*Q*), *and using the result in Theorem 2, we obtain that with probability* 1 − *o*_*p*_(1),

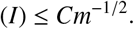

*For term (II), we rely on the following bounds:*

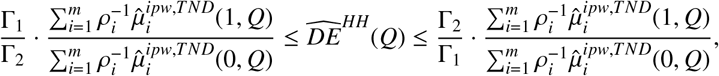

*where the right-hand side is fully estimable given* (Γ_1_, Γ_2_).

*For term (III), we analyze the difference between sample averages and population expectations. Fixing v* ∈ {0, 1}, *define the oracle version of the IPW estimator as* 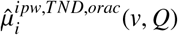 *by replacing* 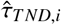 *with the true τ*_*T ND,i*_. *We then decompose*

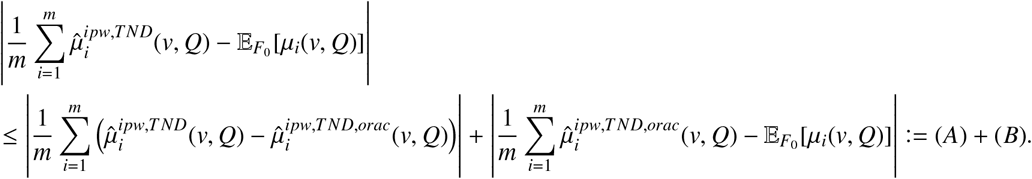

*For term (A), we have:*

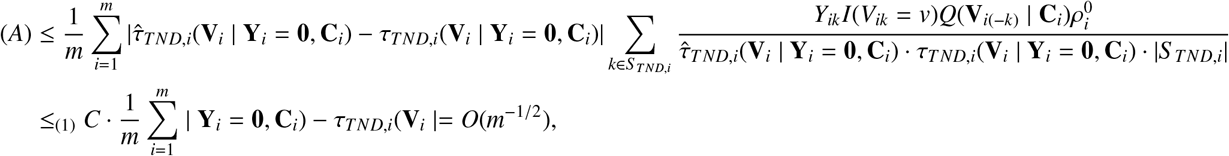

*where (1) uses Corollary 1 and the boundedness of all terms in the summand. For term (B), we apply Chebyshev’s inequality:*

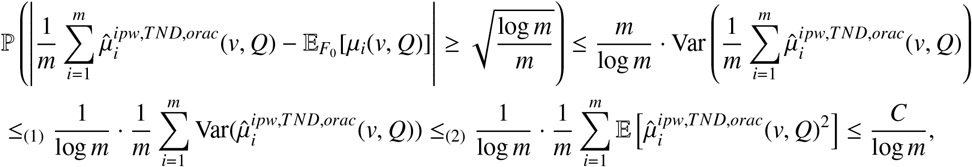

*where (1) uses the independence of clusters, and (2) follows from boundedness of the outcomes, policies Q and τ*_*T ND,i*_. *Combining bounds for terms (I), (II), and (III) completes the proof*.

## 4 Conclusion

In this work, we developed a methodological framework for estimating vaccine effectiveness under the test-negative design while accounting for partial interference. By defining policy-relevant direct and spillover effects using the geometric mean of risk ratios, we introduced inverse-probability weighted (IPW) estimators that remain valid under TND sampling. Our theoretical results establish the identifiability and large-sample properties of these estimators, ensuring their consistency and asymptomatic normality.

## Data Availability

All data produced in the present work are contained in the manuscript

## Acknowledgments

The authors gratefully acknowledge a Canadian Institutes of Health Research Project Grant, and CJ is grateful for the postdoctoral fellowship awarded by the StatLab at the Centre de Recherches Mathématiques. DT is supported by a research-career award from the Fonds de recherche du Québec – Santé. MES holds a Canada Research Chair in Causal Inference and Machine Learning in Health Science.

## SUPPLEMENTARY MATERIAL FOR

### Estimating COVID-19 Vaccine Effectiveness with Partial Interference under Test-Negative Design Sampling

#### Description

This supplementary material includes the proofs of the theoretical results and additional simulation studies. All the code for conducting simulation studies and analyzing real data is accessible in the online repository https://github.com/CONGJIANG/TNDDR.

## A Proof of Identification

With a length *N*_*i*_ row vector of ones 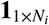 and column vector 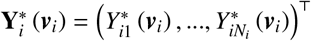, we have

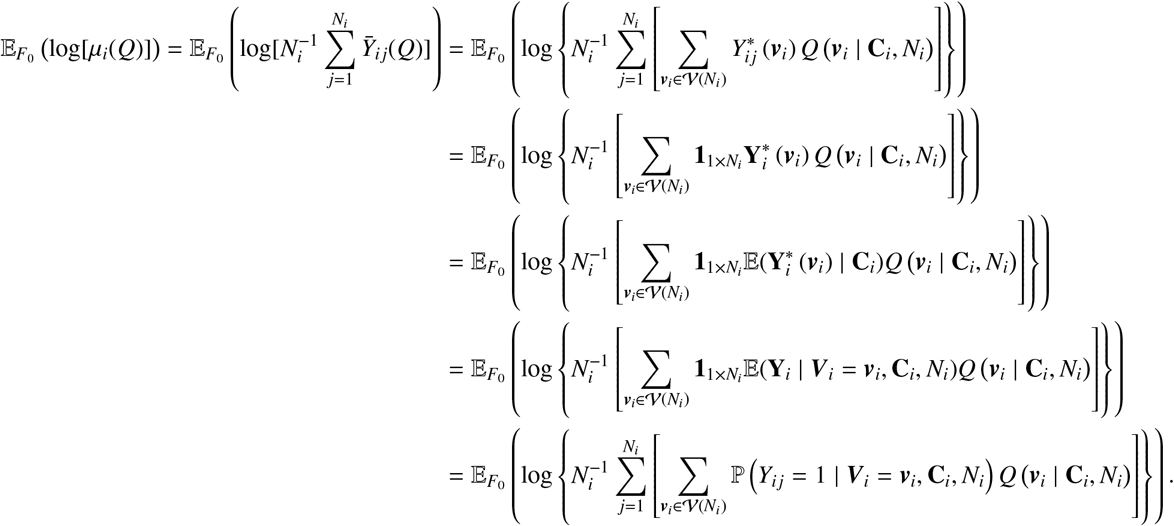

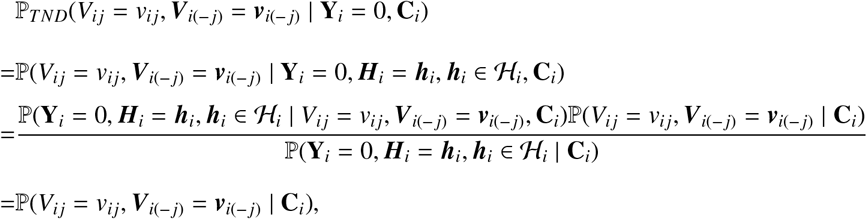

where the fourth equation follows from iterated expectation, and the fifth equation from conditional exchangeability and causal consistency.

## B Proof of Lemma 2 and Theorem 1

### B.1 Proof of Lemma 2

We want to show that the cluster-level propensity score *τ*_*i*_ = ℙ (*V*_*i j*_ = *v*, ***V***_*i*(− *j*)_ = ***v***_*i*(− *j*)_ | **C**_*i*_) can be identified as ℙ _*T ND*_( *V*_*i j*_ = *v*, ***V***_*i*(− *j*)_ = ***v***_*i*(− *j*)_ | **Y**_*i*_ = 0, **C**_*i*_), under the cluster-level control exchangeability assumption, i.e., {**Y**_*i*_ = 0, ***H***_*i*_ = ***h***_*i*_, ***h***_*i*_ ∈ ℋ_*i*_} ⫫ ***V***_*i*_ | **C**_*i*_.

Cluster-level control exchangeability assumption: being hospitalized for symptoms of *another infection* is independent of vaccination conditional on covariates, that is {**Y**_*i*_ = 0, ***H***_*i*_ = ***h***_*i*_, ***h***_*i*_ ∈ ℋ_*i*_} ⫫ ***V***_*i*_ | **C**_*i*_, or 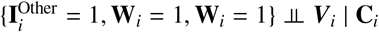.

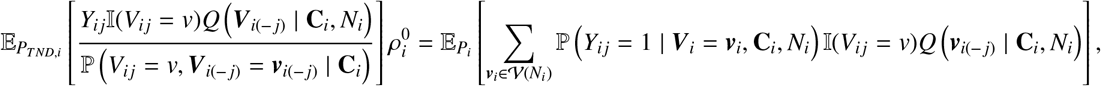

where the cluster-level control exchangeability assumption implies the last equation based on the fact that ℙ (**Y**_*i*_ = 0, ***H***_*i*_ = ***h***_*i*_, ***h***_*i*_ ∈ ℋ_*i*_ | *V*_*i j*_ = *v*_*i j*_, ***V***_*i*(− *j*)_ = ***v***_*i*(− *j*)_, **C**_*i*_) = ℙ (**Y**_*i*_ = 0, ***H***_*i*_ = ***h***_*i*_, ***h***_*i*_ ∈ ℋ_*i*_ | **C**_*i*_).

### B.2 Proof of Theorem 1

In this section of the appendix, we prove Theorem 1 by focusing primarily on establishing *the TND identification equality*, which involves Lemma 2 and another lemma (i.e., Lemma 5) which is presented in the following proof.

**Proof:** Under the assumptions that (1) ℙ_*i*_(**Y**_*i*_ = **y**_*i*_ | ***V***_*i*_, **C**_*i*_, ***H***_*i*_ = ***h***_*i*_, ***h***_*i*_ ∈ ℋ_*i*_) = 0 for **y**_*i*_ ∈ {**0, 1**}; (2) 0 < ℙ_*i*_ ***V***_*i*(− *j*)_ = ***v***_*i*(− *j*)_ | **C**_*i*_, ***H***_*i*_ = ***h***_*i*_, ***h***_*i*_ ∈ ℋ_*i*_ < 1; (3) 0 < ℙ_*i*_(**C**_*i*_ = **c**_*i*_ | ***H***_*i*_ = ***h***_*i*_, ***h***_*i*_ ∈ ℋ_*i*_) < 1, we want to show the *TND identification equality*, that is,

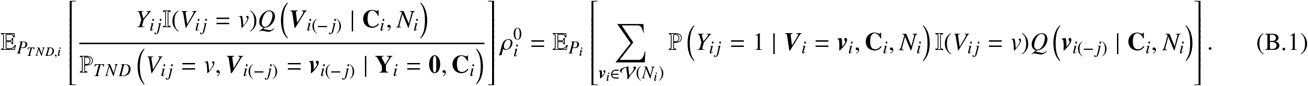

First, in Lemma 5, we show that

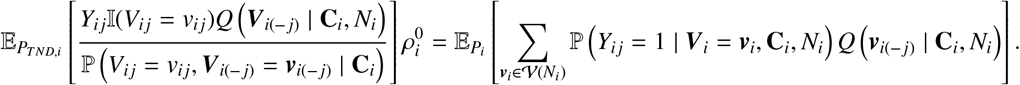

to demonstrate the unbiasedness of 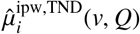 when the cluster-level propensity score *τ*_*i*_ = ℙ (*V*_*i j*_ = *v*, ***V***_*i*(− *j*)_ = ***v***_*i*(− *j*)_ | **C**_*i*_) is known.

#### Lemma 5

*If the cluster-level propensity score τ*_*i*_ = P *V*_*i j*_ = *v*, ***V***_*i*(− *j*)_ = ***v***_*i*(− *j*)_ | **C**_*i*_ *is known, then we have*

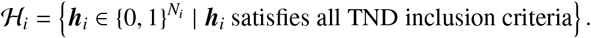

**Proof:** Note that for cluster *i*, we let ***H***_*i*_ = (*H*_*i*1_, …, *H*_*i j*_, …, *H*_*iN*_*i*)^⊤^ be the TND inclusion indicator vector for cluster *i*, and denote the TND inclusion probability for cluster *i* as 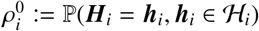, where ℋ_*i*_ denotes the set of valid inclusion patterns for the TND sample in cluster *i*, i.e.,

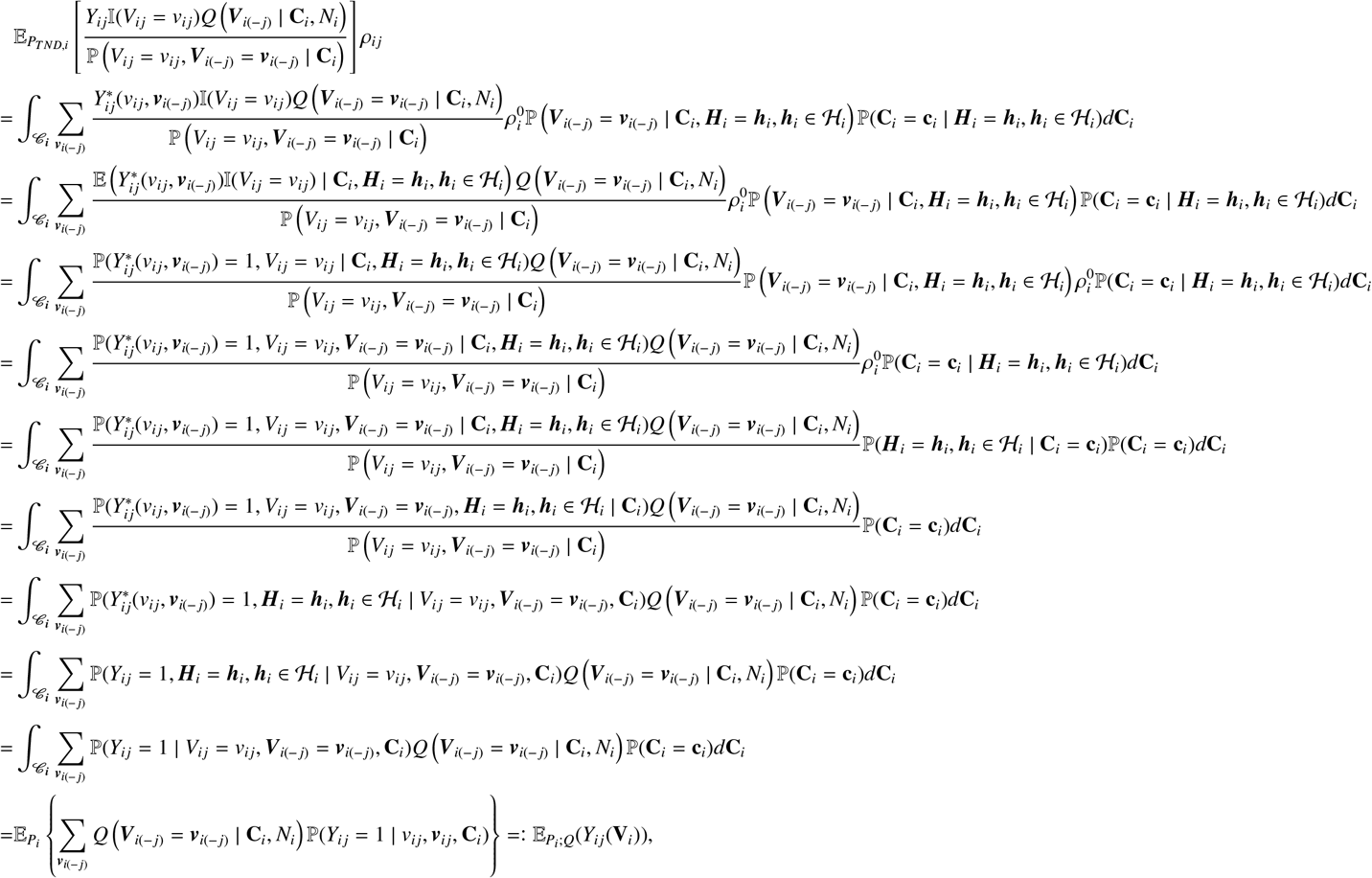

Let 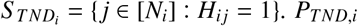 to be the distribution of (**Y**_*S*_ *TND,i* (**v**_*i*_), **V**_*S*_ *TND,i*, **C**_*S*_ *TND,i*). *P*_*i*_ is the distribution of (*Y*_*i*_(**v**_*i*_), **V**_*i*_, **C**_*i*_) and *ρ*_*i j*_ := ℙ (*H*_*i j*_ = 1). Then for each unit *j* in cluster *i*, we have

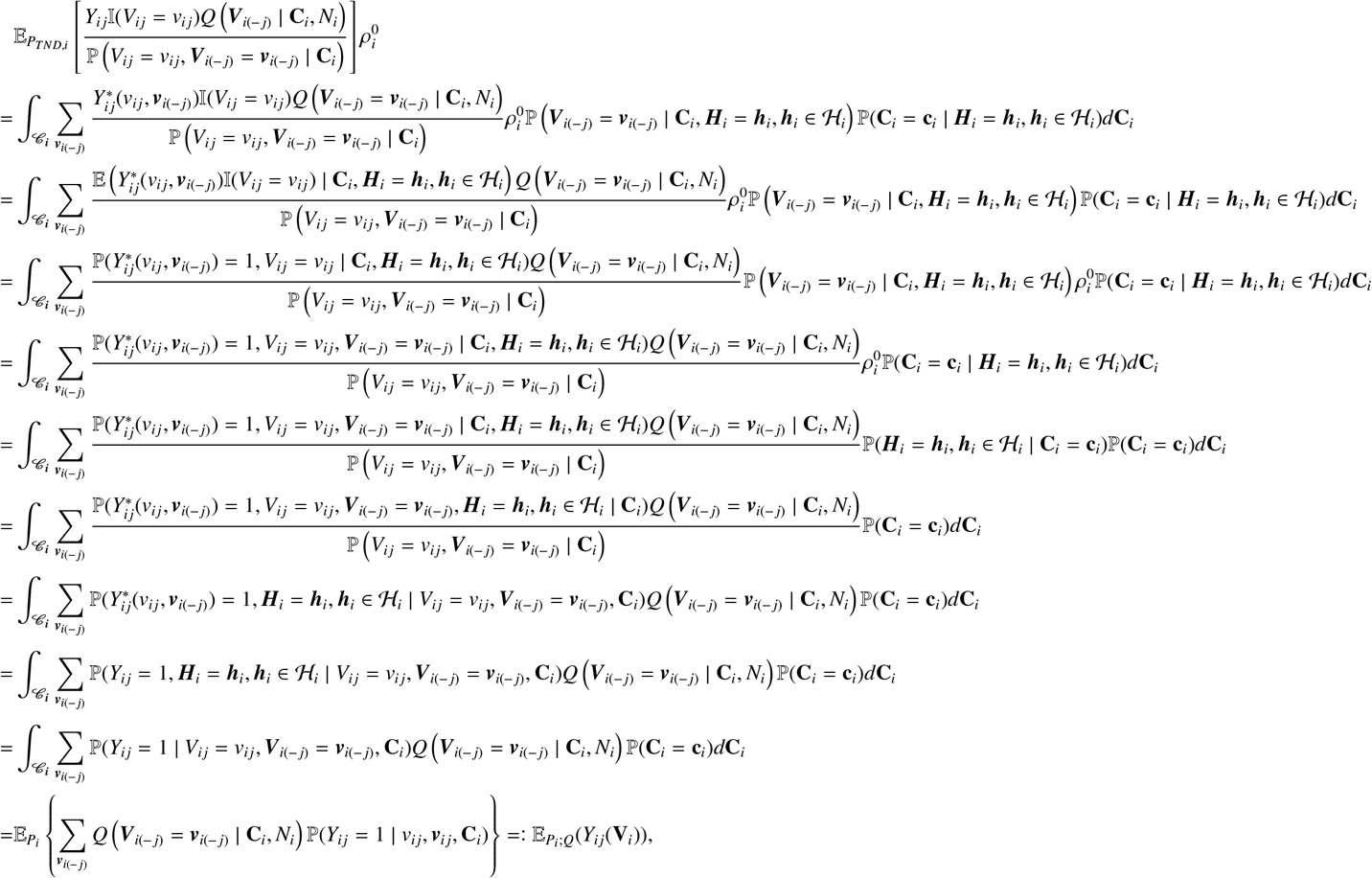

where the first equality follows by the definition of 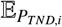 (w.r.t. distribution ℙ (**C**_*i*_ = **c**_*i*_ | ***H***_*i*_ = ***h***_*i*_, ***h***_*i*_ ∈ ℋ_*i*_)), the law of total expectations (w.r.t. distribution ℙ (***V***_*i*(− *j*)_ = ***v***_*i*(− *j*)_ | **C**_*i*_, ***H***_*i*_ = ***h***_*i*_, ***h***_*i*_ ∈ ℋ_*i*_) and the causal consistency assumption that 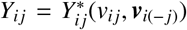; the second equality is by the law of total expectation. The fourth equality follows by the conditional exchangeability or ignorability assumption that 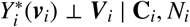; the fifth equality follows by the Bayes rule that 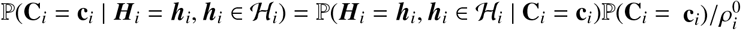.

Therefore, we have

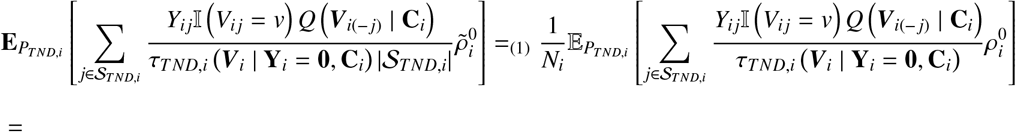

- If bias sampling is on cluster level, then *ρ* is identical across different clusters. All asymptotics works. The problem is for bias cancellation. Assume 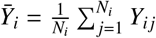, then the bias cancelling can happen is we assume 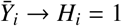. But notice that this is a sufficient condition, not the necessary one, meaning it is still possible that 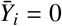 and *H*_*i*_ = 1.
- Problem is on how average of TND sample converges to the expectation of its expectation. If using law of large number, the independent should appear in some extent. If assume neighborhood interference, we don’t know full network information. If assume the hypothetical and realized treatmtent the same, more plausible on experiment design, but not plausible on the observational studies where the realized one always is not the one we want to learn.

Regarding the inverse probability weighting estimators under TND (i.e., Equation 16):

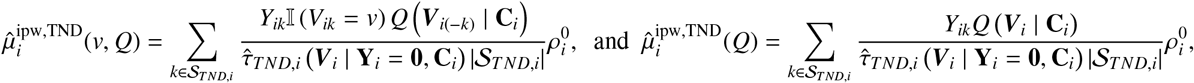

we have the unbiasedness result in the theorem, which follows directly from the result of Lemma 5.

## C Hájek-type IPW esitmators

In this section, we are presenting the Hájek-type IPW estimators for µ_*i*_(*v, Q*) and µ_*i*_(*Q*). With correctly-specified estimated cluster-level propensity score under TND sampling,

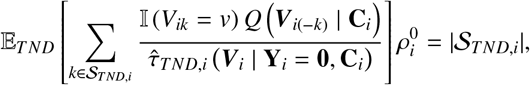

so that replacing |𝒮_*T ND i*_| by its consistent estimator 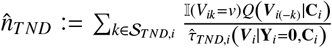 yields the Hájek-type (Hájek (1971); Liu et al. (2016)) IPW esitmator for µ_*i*_(*v, Q*):

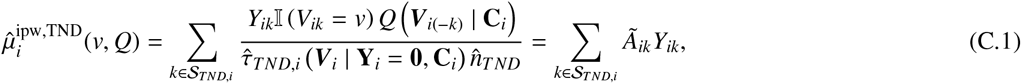

Where

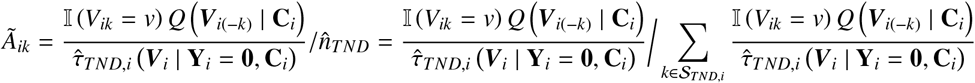

and 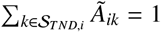Similarly, the Hájek-type IPW estimator for µ_*i*_(*Q*) takes the form

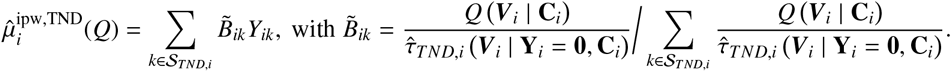

Notably, the Hájek-type IPW estimator demonstrates superior efficiency compared to the traditional IPW estimator when applied to finite samples Särndal et al. (2003).

## D Proof of Theorem 2

This section presents the proof of Theorem 2. We begin by examining the estimation equations derived from both the cluster-level propensity score (referenced in Equation 17) and those used to solve the target effects (e.g., Equation 20).

Firstly, we focus on the estimation equations associated with the cluster-level propensity score:

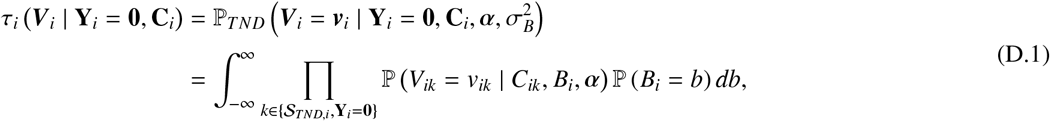

Where 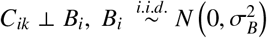, and P (*V*_*ik*_ = 1 | *C*_*ik*_, *B*_*i*_, *α* = *g*^−1^ *α*^⊤^*C*_*ik*_ + *B*_*i*_). Hence, the log-likelihood for the mixed effects model is written as 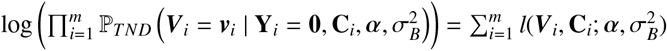, where

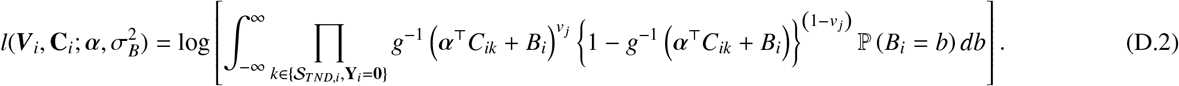

For the score function system that is 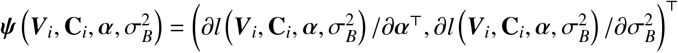 of Equation 21, we denote each element in 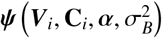 as

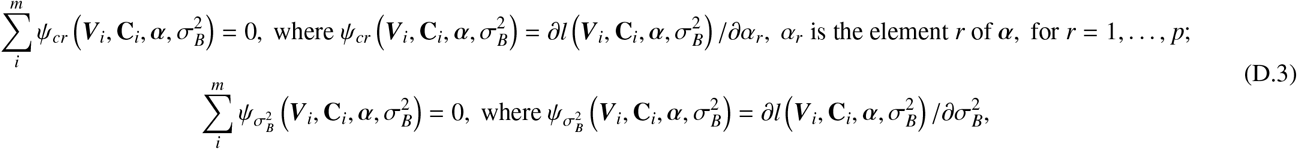

and 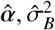 that maximize the log-likelihood are the solutions.

As mentioned in the main context, it is important to note that 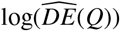 is a solution for log(*DE*(*Q*)) to the estimating equation

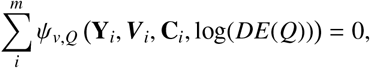

where

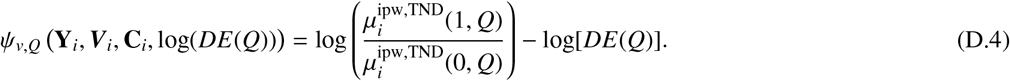

Therefore, with parameters 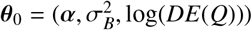, we have the estimator, 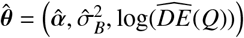 which is a solution to the estimating equation systems ∑_*i*_ *ψ* (**O**_*i*_, *θ*) = **0**, with **O**_*i*_ = **Y**_*i*_, ***V***_*i*_, **C**_*i*_, where

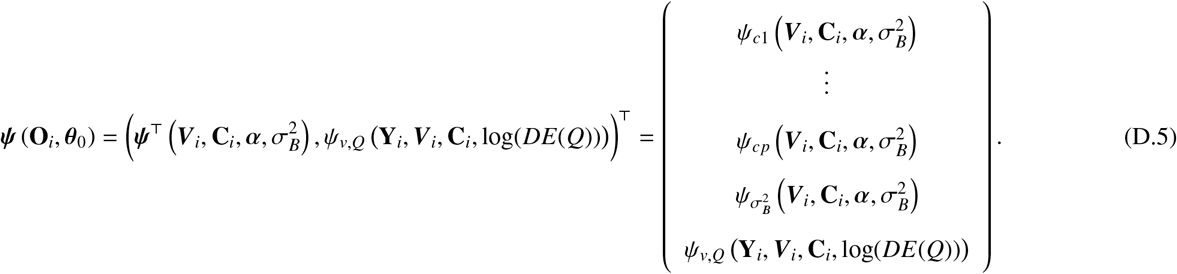

Hence, according to M-estimation theory (Boos et al. (2013)), 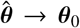 in probability, and the difference 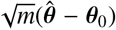 converges in distribution to a multivariate normal distribution *N*(**0**, Σ), where the variance matrix Σ adopts a sandwich form of

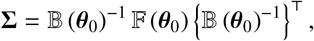

Where

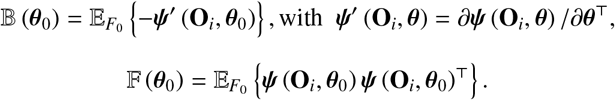

Note that *θ*_0_ is the true parameter value defined Stefanski and Boos (2002) by

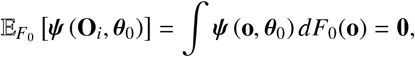

where here *F*_0_ denotes the cumulative distribution function of **O**_*i*_.

In the subsequent sections, we present the specific formulation for the asymptotic variance of 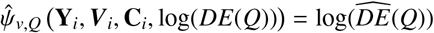, explicitly expressing 𝔹 (*θ*_0_) and 𝔽 (*θ*_0_) in terms of *ψ*^′^(**O**_*i*_, *θ*) and *ψ*(**O**_*i*_, *θ*). First, *ψ*^′^ (**O**_*i*_, *θ*) is the (*p* + 2) × (*p* + 2) matrix:

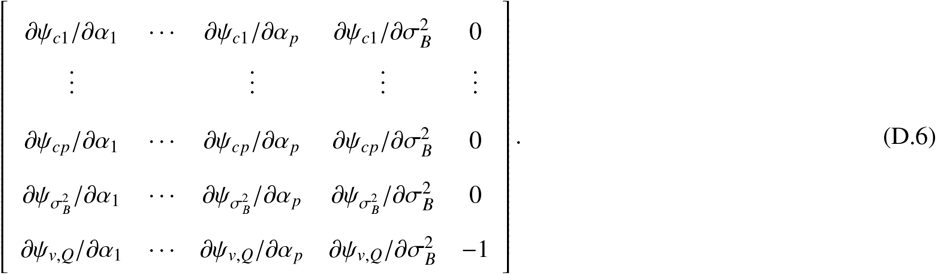

Then, using block matrix notation, 𝔹 (*θ*_0_) can be written as

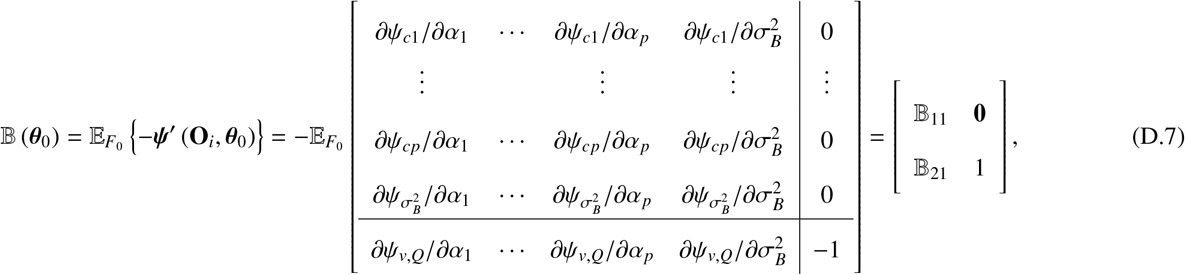

where 𝔹_11_ is a (*p* + 1) × (*p* + 1) information matrix of the cluster PS, and 𝔹_21_ is a 1 × (*p* + 1) vector. F (*θ*_0_) can be written as

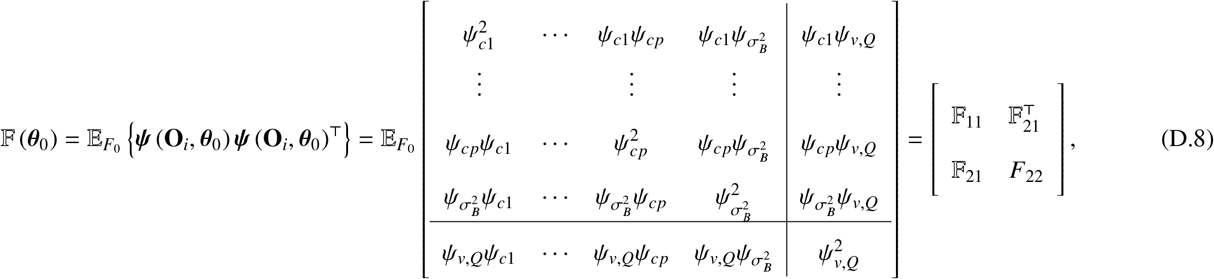

where 𝔽_11_ is a (*p* + 1) × (*p* + 1) matrix, 𝔽_21_ is a 1 × (*p* + 1) vector, and *F*_22_ is a scalar. In addition,

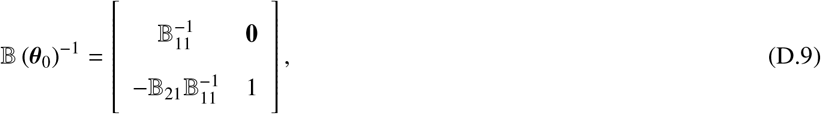

and the variance matrix Σ can be written as

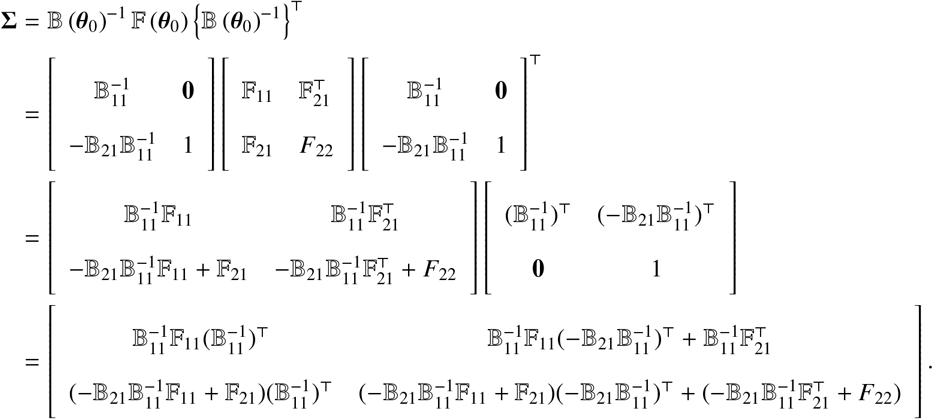

As 𝔹_11_ and 𝔽_11_ are associated with the score equations of the log-likelihood function in the mixed effects model, it can be inferred that 𝔽_11_ is equal to F_11_ (Stefanski and Boos (2002)). Then, we can simplify the matrix Σ:

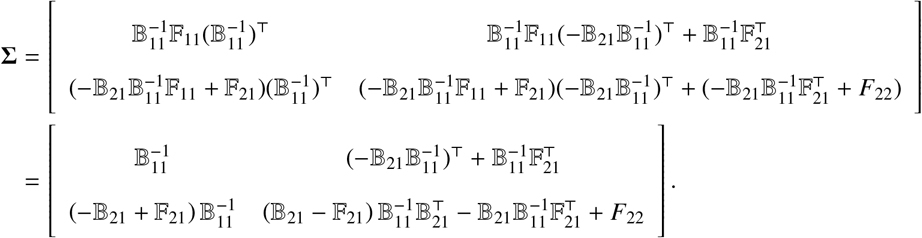

Therefore, we conclude that expressed as 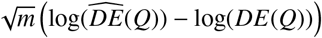 converges in distribution to *N*(0, Σ_*log*(*DE*)_), as *m* → ∞, where Σ_*log*(*DE*)_ is expressed as

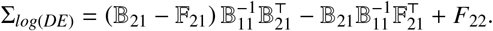

Further, applying the delta method with the exponential function such that exp[log(*DE*(*Q*))] = *DE*(*Q*), we can conclude that 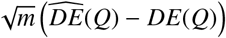 converges in distribution to *N*(0, Σ_*DE*_), as *m* → ∞, where Σ_*DE*_ = (*DE*(*Q*))^2^Σ_*log*(*DE*)_.

By replacing 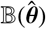 and 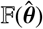 with their empirical estimators, we obtain:

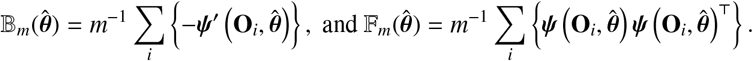

Thus, the empirical sandwich variance estimator is given by: 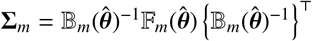. The element in the bottom-right position, i.e., (*p* + 2, *p* + 2) of the matrix Σ_*m*_, when multiplied by *m*^−1^, provides the estimated asymptotic variance for 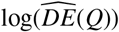 and thus 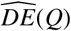.

In conclusion, this implies the estimation of the asymptotic variance of 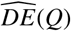 by

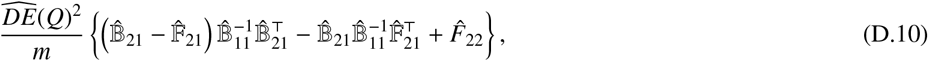

where 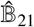 represents the first *p* + 1 terms in the bottom row of 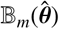, and 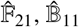, and *F*_22_ are the corresponding sub matrices or elements of 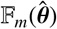.

### Remark 1

For further computation of variance of DE, we note that the explicit form of 𝔹_21_ and 𝔽_21_ can be derived as follows. First, regarding 𝔹_21_, we have

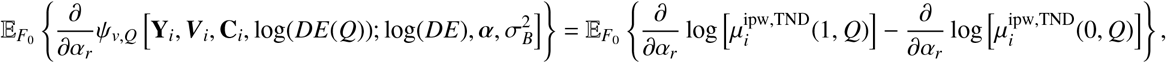

where for *v* = {0, 1},

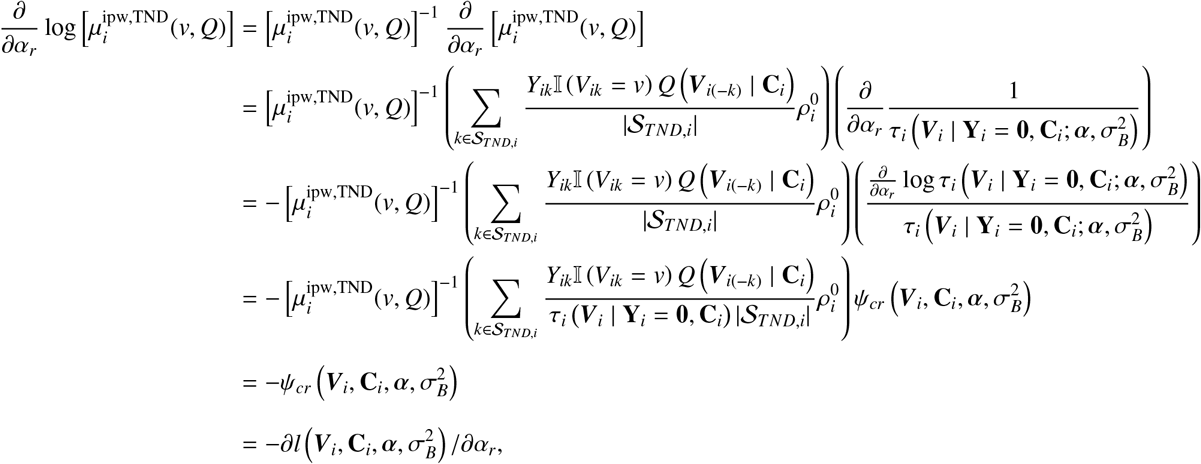

Where 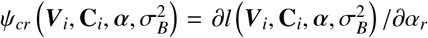, the score function of the cluster PS, related to the PS log-likelihood for the mixed effects model (D.2). Similarly, a corresponding process can be derived for the parameter 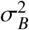 to complete the derivation of the 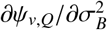 element in 𝔹_21_:

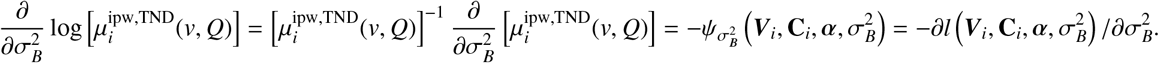

Then, for 𝔽_21_, with the score function of the cluster 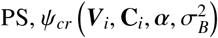, we have

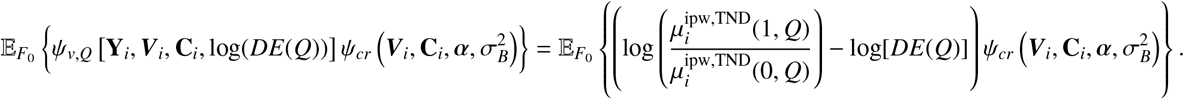

### Remark 2

The estimation of large-sample variances for indirect, total, and overall effect estimators follows a similar approach, with different estimation equations that:

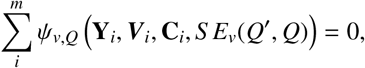

Where

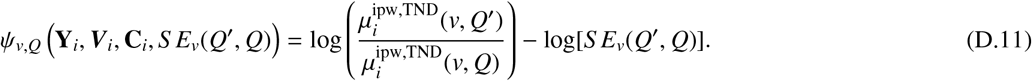

For the total and overall effect, we have that

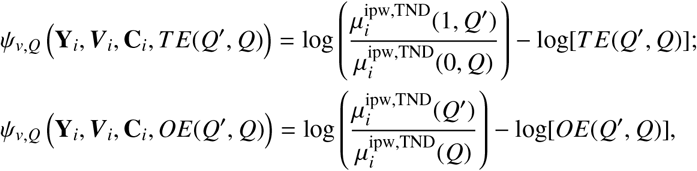

where, again,

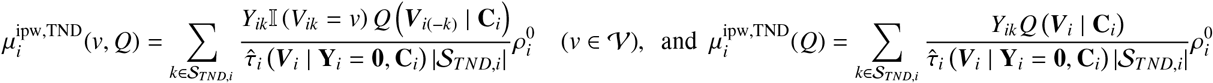

and

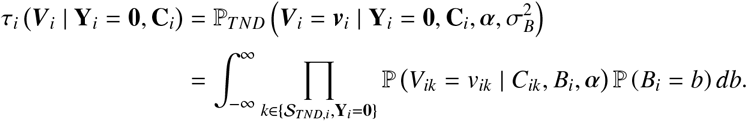

Specifically, this involves substituting Equation (D.4) with the target estimation equation (e.g., for SE, ψ_*v,Q*_ (**Y**_*i*_, ***V***_*i*_, **C**_*i*_, *S E*_*v*_(*Q*^′^, *Q*))) in the estimation equations from the system (D.5) and subsequently computing the corresponding 𝔹 (*θ*_0_) and 𝔽 (*θ*_0_) based on *ψ*^′^(**O**_*i*_, *θ*) and *ψ*(**O**_*i*_, *θ*). For instance, for the computation of IE, we derive the explicit form of 𝔹_21_ and 𝔽_21_. First, regarding B_21_, we have

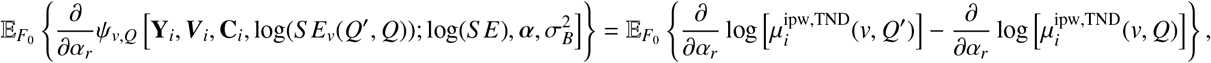

where for *v* = {0, 1},

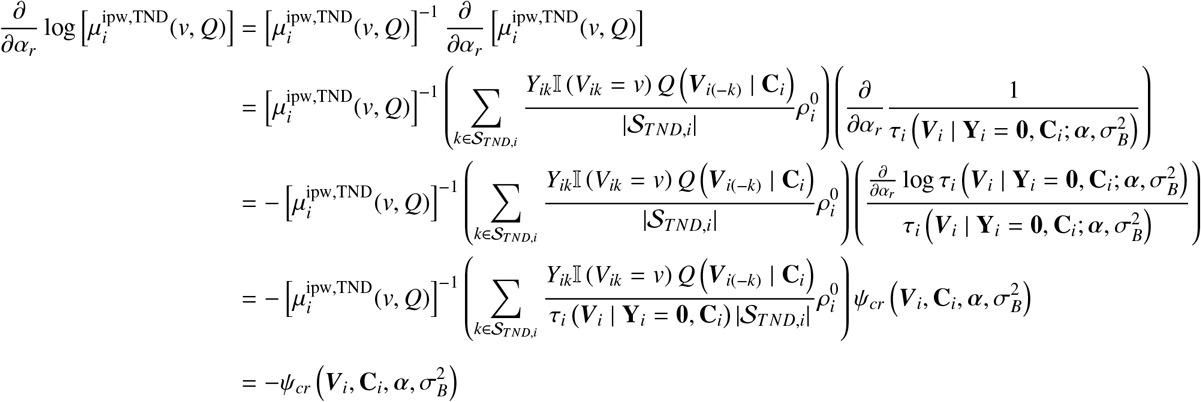

Where 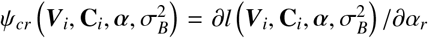, the score function of the cluster PS, related to the PS log-likelihood Equation D.2.

Similarly, a corresponding process can be derived for the parameter 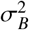 to complete the derivation of the 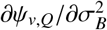 element in 𝔹_21_.

Then, for 𝔽_21_, with the score function of the cluster 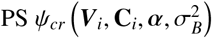, we have

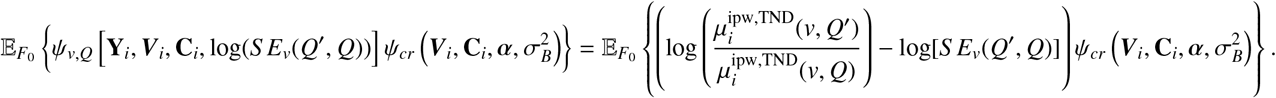

- 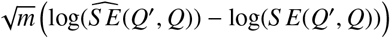 converges in distribution to *N*(0, Σ_*log*(*S E*)_), as *m* → ∞, where Σ_*log*(*S E*)_ is expressed as

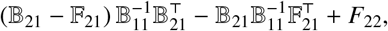

Where

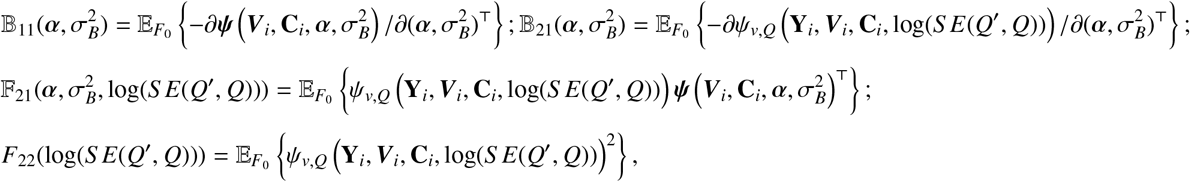

regarding estimation equations in Equations D.11 and 21.

## E Proof of Lemmas 1 and 4

**Proof 3 (Proof of Lemma 1)** *Let function* 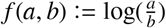. *By taking Taylor expansion on* (𝔼 (*a*), 𝔼 (*b*)) *up to the second order of f* (*a, b*), *we have*

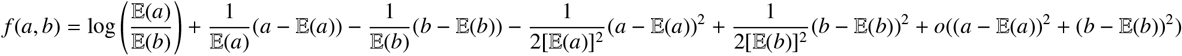

*Therefore*, 𝔼 (*f* (*a, b*)) *satisfies*

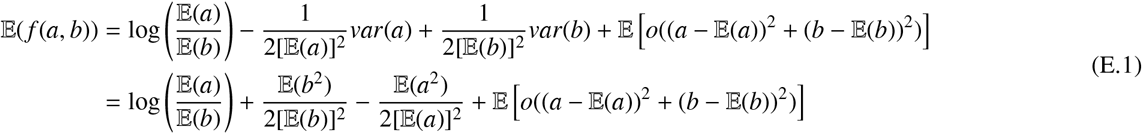

*Let a* = µ_*i*_(1, *Q*) *and b* = µ_*i*_(0, *Q*), *we then have*

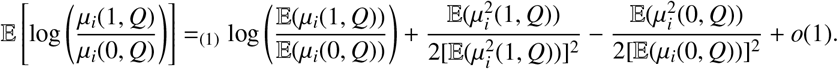

- *follows from Theorem 1, together with the fact that* µ_*i*_(1, *Q*) *and* µ_*i*_(0, *Q*) *are bounded. Therefore, we have*

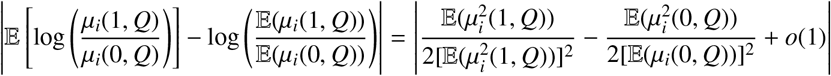

*By utilizing mean value theorem on function* exp(*x*), *we have*

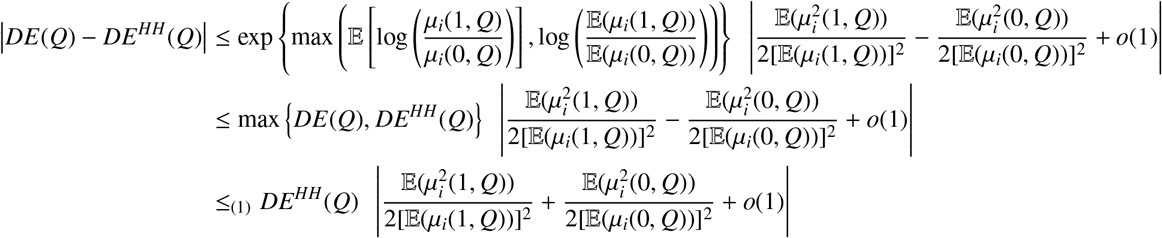

*(1) is by Jensen’s inequality and DE*(*Q*), *DE*^*HH*^(*Q*) ≥ 0. **Proof 4 (Proof of Lemma 4)** *Based on Lemma 1, we have*

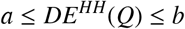

*where* 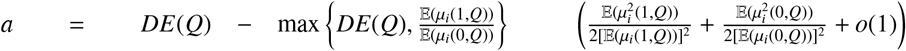 *and* 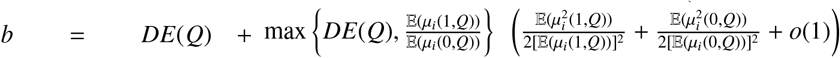. *A consistent estimator of b and its upper bound are*

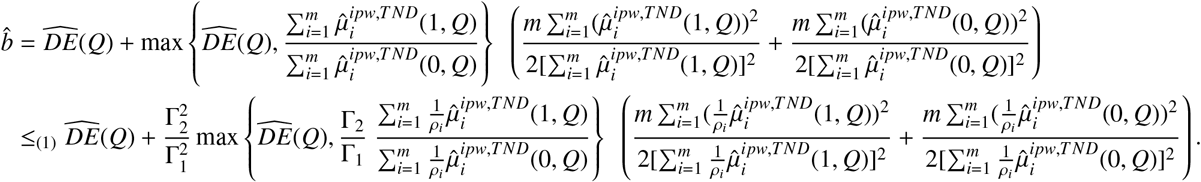
- *is based on taking the upper bound* Γ_2_ *of ρ*_*i*_ *in the numerator and the lower bound of* Γ_1_ *of ρ*_*i*_ *in the denomenator and each estimator is larger or equal to* 0. *Similarly, a consistent estimator of a and its lower bound are*

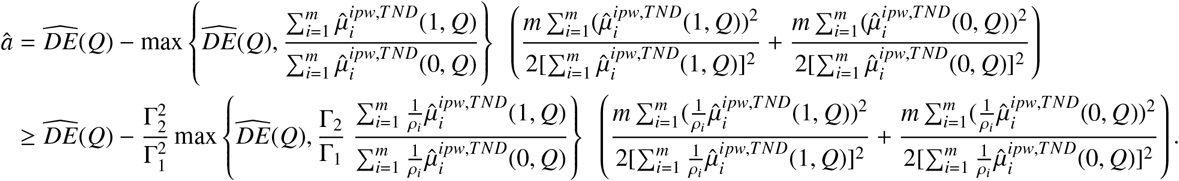

*Therefore, we have, with probability* 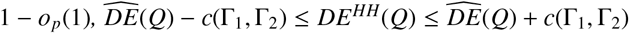 *where*

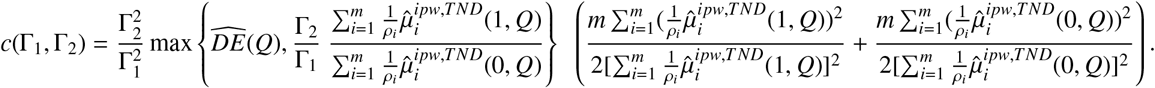

